# Screening for patients at risk for cardiac amyloidosis via electronic health records: A multicenter machine learning development and validation study

**DOI:** 10.64898/2026.04.27.26351820

**Authors:** Clemens P. Spielvogel, David Kersting, David Haberl, Maximilian Autherith, Laurenz Hauptmann, Josef Yu, Juliane Hennenberg, Kilian Kluge, Kim Moon, Stephan Settelmeier, Jing Ning, Katarina Kumpf, Markus Köfler, Felix Hofer, Katharina Mascherbauer, Julia Mascherbauer, Andreas Kammerlander, Tatjana Traub-Weidinger, Gregor Kasprian, Tienush Rassaf, Jens Kleesiek, Ken Herrmann, Philipp Bartko, Christian Hengstenberg, Marcus Hacker, Raffaella Calabretta, Christian Nitsche

**Author notes:** Correspondence to: Christian Nitsche, MD, PhD, Division of Cardiology, Department of Internal Medicine II Medical University of Vienna, Spitalgasse 23, A-1090 Vienna/Austria.

## Abstract

**Background:** Timely detection is crucial to improve outcomes in patients with cardiac amyloidosis (CA) by initiation of life-saving treatments. Although confirmatory bone scintigraphy is highly accurate for CA detection, identifying at-risk patients for referral remains challenging.

**Objectives:** This study aimed to develop and validate a machine learning model, *Amylo-Detect*, using structured multimodal electronic health record (EHR) data to guide referrals for confirmatory scintigraphy and monoclonal protein testing.

**Methods:** Consecutive all-comer patients (n=11,616) referred for bone scintigraphy at the Vienna General Hospital (2010-2023) were retrospectively included. Patients referred before August 2020 formed the development cohort. The remaining patients comprised the internal validation cohort. External validation was performed at the University Hospital Essen (n=1,521). *Amylo-Detect* was trained using 50 routinely available parameters to predict CA-suggestive uptake (Perugini grade ≥2) and compared with an existing score and clinical routine.

**Results:** High-grade uptake was present in 388 patients (3.0%). *Amylo-Detect* demonstrated excellent performance in development (AUC 0.93), independent internal validation (AUC 0.91), and external validation cohort (AUC 0.91), outperforming existing scoring systems and clinical routine. Results were consistent across subgroups, even when crucial predictors were missing. Of the 42/388 (10.8%) patients missed in clinical routine, 12/42 (29%) were additionally detected by *Amylo-Detect*. The model further conveyed significant prognostic value for mortality and heart failure hospitalization.

**Conclusions:** We present *Amylo-Detect*, a validated EHR-based tool for CA risk prediction, available as a web app, allowing application and further evaluation. By improving timely detection and referral, *Amylo-Detect* promises to address diagnostic delays and improve outcomes.

**Author summary:** Cardiac amyloidosis is a progressive and often fatal heart disease that is frequently diagnosed too late, even though effective treatments are now available. A major challenge is recognizing which patients should be referred for specialized testing, because early symptoms are diverse and often non-specific. In this study, we developed and validated a machine learning prediction system, called Amylo-Detect, that uses routinely collected information from electronic health records to identify patients who may be at risk for cardiac amyloidosis. We trained and externally validated the tool using data from 13,137 patients across two hospitals. We found that Amylo-Detect was highly accurate in identifying patients with disease-indicative findings on confirmatory scans and identified a substantial number of patients who were not initially suspected of having cardiac amyloidosis. Amylo-Detect, which we make publicly available via a web app, consistently outperformed existing risk scores and routine clinical decision-making. Our findings suggest that automated analysis of electronic health records can support clinicians in recognizing cardiac amyloidosis earlier, reduce diagnostic delays, and potentially improve patient outcomes by enabling timely treatment.

## Introduction

Cardiac amyloidosis (CA) is a progressive and fatal disease characterized by misfolded proteins aggregating to amyloid fibrils with subsequent deposition in the myocardium, leading to heart failure and death. The two major types of CA include transthyretin (ATTR) and light chain (AL) amyloidosis, making up for more than 98% of currently diagnosed CA cases^1^. Over the last decade ATTR-CA in particular has emerged as substantially more common than previously appreciated, accounting for approximately 6-13% of patients with heart failure with preserved ejection fraction^2,3^, 13% of elderly patients with severe aortic stenosis^4^, and 1.5% (Perugini grade≥2) to 3.3% (Perugini grade≥1) among all-comer patients undergoing bone scintigraphy^5^. However, despite considerable advances regarding disease awareness, CA is still considered underdiagnosed and the early and accurate identification of patients remains a major challenge, particularly as disease-modifying treatments are believed to be most effective in early disease stages^6–8^. A critical bottleneck is raising the initial suspicion of CA early and accurately to refer patients to confirmatory testing by scintigraphy and monoclonal protein testing.

Amyloidosis is a systemic disease and can manifest through a diverse array of symptoms such as neuropathy, renal dysfunction, and gastrointestinal issues. These indicators are non-specific for CA when observed in isolation. However, the presence of multiple indicators can help to raise an initial suspicion. This multi-symptomatic presentation makes CA an ideal target for data-driven, multimodal approaches that can integrate routinely collected parameters and uncover complex, non-linear patterns that predict CA.

Prior artificial intelligence (AI) efforts in amyloidosis have shown promise when applied to cohorts with a high pretest probability.^9,10^ However, such methods typically do not generalize when deployed across all-comer patients where the prevalence is low and the clinical presentation heterogeneous, limiting their applicability in clinical practice.

In this study, we present *Amylo-Detect*, a fully automated machine learning tool developed and validated using routinely available pre-scintigraphy parameters from electronic health records (EHR) of consecutive, all-comer patients referred for bone scintigraphy and independently validated at two sites. *Amylo-Detect* predicts CA-suggestive tracer uptake (Perugini grade ≥2) to guide referral for confirmatory testing. This approach aims to close diagnostic gaps in CA by accelerating the identification of at-risk patients, enabling timely confirmatory testing and treatment to ultimately improve cardiovascular outcomes.

## Methods

### Study design and participants

This large-scale multicenter cohort study included consecutive all-comer referrals for whole-body ^99m^Tc-3,3-diphosphono-1,2-propanodicarboxylic acid (DPD) scintigraphy at the Vienna General Hospital (Austria) between January 2010 and February 2023. Patients examined before July 2020 were included for development and the remaining patients for independent internal validation (**Supplementary Table S1 and S2**). For external validation, all-comer referrals for DPD scintigraphy were collected at the University Hospital Essen (Germany), acquired between August 2018 and April 2025 (**Supplementary Table S3**). Beyond scintigraphy images, collected data included corresponding baseline parameters from EHR, including demographic, blood, echocardiography, and comorbidities. For external validation, the six most important parameters were collected to enable model inference. To reflect a realistic clinical routine scenario, patients with missing values in baseline parameters were retained to ensure clinical applicability. Tricuspid, pulmonary, mitral, and aortic valve parameters included stenosis as well as regurgitation and were dichotomized into moderate or higher and less than moderate. All-cause mortality and heart failure-associated hospitalization (HFH) were assessed in the Vienna cohorts and were 100% and 92% complete, respectively. All-cause mortality was captured via the Austrian Death Registry. HFH was determined from patient records of the Vienna General Hospital, the Vienna-Health-Association database, and the nationwide electronic health records.

### Diagnostic model for prediction of CA-suggestive uptake

Machine learning was performed to predict high-grade cardiac uptake on scintigraphy indicative of CA using the 50 baseline parameters. The machine learning process consisted of model development and cross-validation, followed by independent internal validation using data from a different time range and external validation. Stratified 10-fold cross-validation was performed on the development cohort. Subsequently, the *Amylo-Detect* model was built on the entire development dataset and validated on the independent internal and external validation datasets. During cross-validation, each fold’s preprocessing included the standardization of features, imputation using k-nearest neighbors, selection of features through the minimum redundancy maximum relevance (mRMR) algorithm^11^, addressing imbalances with the synthetic minority oversampling technique (SMOTE)^12^, and automated hyperparameter tuning via a random search within a pre-defined hyperparameter grid. Preprocessing algorithms were applied in the aforementioned order. Any kind of fitting procedures during the entire modelling process, including preprocessing steps, were solely performed on the training data to avoid any kind of data leakage. Training was performed using seven machine learning algorithms, including random forest, k-nearest neighbors, extreme gradient boosting, logistic regression, support vector machine, decision tree, and explainable boosting machine. Risk calibration was performed using isotonic regression. For binary classification, a probability cut-off of 0.77 was determined in the development data and employed to ensure a reasonable balance between diagnostic metrics. However, by outputting a continuous probability score, *Amylo-Detect* allows clinicians the flexibility to interpret or adjust the threshold based on the clinical context. All fitted models, including standardization, imputation, feature selection, hyperparameter tuning, classification modelling, and calibration, were fitted during training and not changed before application to any validation cohorts. The entire model, including all preprocessing, is publicly available via the free *Amylo-Detect* Web App (https://amylo-detect.streamlit.app/).

### Comparison with existing risk score and clinical routine

*Amylo-Detect* was evaluated against the Mayo ATTR-CM score, which has previously demonstrated strong performance in both a pyrophosphate (PYP) scan referral cohort and in patients with heart failure with preserved ejection fraction (HFpEF)^9^. The comparison was conducted in the all-comer cohort and a small subgroup with high pre-test probability, defined as an extracellular volume (ECV) >40% on cardiac magnetic resonance imaging (CMR)^13–15^.

For the comparison of *Amylo-Detect* with clinical routine, referral indications were collected for all scans. *Amylo-Detect* was compared with the clinical referral indication with respect to the same diagnostic performance metrics based on whether the referred patient was found to have high-grade cardiac uptake.

### Statistical analysis

Continuous parameters are reported as mean ± standard deviation (SD) or median with interquartile range (IQR). Categorical data are presented as counts and percentages. To compare patient characteristics across groups, numerical variables were analyzed using Mann-Whitney U test or T-test, and categorical variables were evaluated either using Fisher’s exact test or Chi-squared test. The Shapiro-Wilk test was applied to determine normality for selecting the appropriate statistical test. The performance of machine learning models is reported using the area under the receiver operating characteristic curve (AUC), and various other diagnostic metrics with corresponding 95% confidence intervals (CIs). CIs were computed over the ten cross-validation folds in the development cohort and using 1,000 bootstrap samples in the independent internal and external validation cohorts. Model AUCs were compared using the DeLong test. If patients underwent multiple scans, only the first scan was employed to avoid any data leakage or model bias. Kaplan-Meier estimates and Cox proportional hazard models were applied to assess association of model predictions with all-cause mortality and HFH, starting with the day of scintigraphy. Multivariable adjustment was performed for history of cancer. *P* values less than 0.05 were deemed statistically significant. All developments and analyses were performed using Python 3.9.5. Prior to all analyses, random seeds were set to the default where available, otherwise to zero. This study and the related reporting adhere to the TRUE-AIM guidelines^16^.

## Results

### Population characteristics

A graphical overview of the study is shown in **Figure 1**. The development cohort included 9,170 patients with a median age of 64 (IQR 52-73) years. Overall, 5,488/9,170 (64%) patients were female. The separately collected internal validation cohort included 2,446 patients with a median age of 67 (IQR 55-76) years and 1,121/2,446 (46%) females. The external validation cohort comprised 1,521 patients with a median age of 69 (IQR 59-79) years and 882/1,521 (58%) females. High-grade cardiac uptake was present in 142/9,170 (1.5%) of the development cohort, in 101/2,446 (4.1%) of the internal validation and in 145/1,521 (9.5%) patients of the external validation cohort. The number of scans performed with the query of suspected CA was 1,003/9,170 (11%) in the development, 576/2,446 (24%) in the internal validation cohort, and 516/1,521 (33.9%) in the external validation cohort. Detailed referral indications for the Vienna cohorts are shown in **Supplemental Figure 1**. Of 388 patients with high-grade cardiac uptake, 42 (10.8%) were referred for non-cardiac indications. Comprehensive patient characteristics of the development, internal, and external validation cohorts are shown in **Supplemental Tables S1**, **S2** and **S3**. A study flow diagram is shown in **Supplemental Figure S2**.

**Figure 1.**
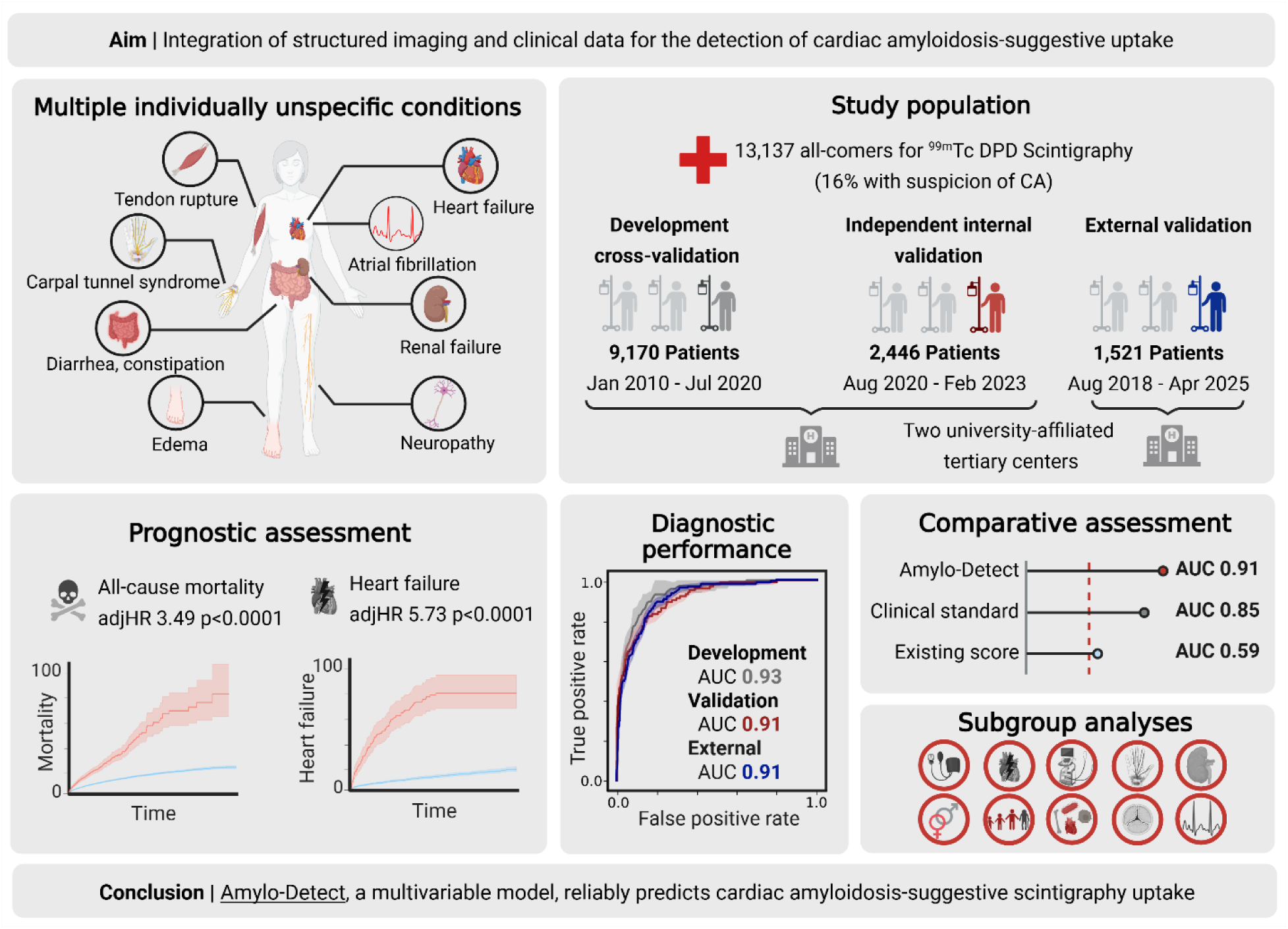
Graphical abstract.

### Diagnostic performance of the machine learning model

During development, the machine learning-enhanced logistic regression achieved the highest performance with a cross-validated AUC of 0.93 (95% CI 0.91; 0.95) (**Supplemental Figure S3**). Consequently, this model, we named *Amylo-Detect*, was employed for all subsequent analyses. In the independent internal validation, *Amylo-Detect* demonstrated robust performance with an AUC of 0.91 (95% CI 0.88; 0.94). Results remained consistent in external validation with an AUC of 0.91 (95% CI 0.89; 0.93). Comprehensive diagnostic metrics are shown in **Table 1**. Initial risk calibration yielded a Brier score of 0.085 and did not improve via isotonic regression (**Supplemental Figure S4**). The six parameters with the highest importance for prediction were (uni- or bilateral) carpal tunnel syndrome followed by atrial fibrillation, sex, diastolic left ventricular function on echocardiography, interventricular septum thickness on echocardiography and patient age.

**Table 1.** Machine learning performance for the prediction of cardiac amyloidosis-indicative uptake on bone scintigraphy.

| Approach | Cohort | AUC<br>(95%<br>CI) | ACC<br>(95%<br>CI) | SNS<br>(95%<br>CI) | SPC<br>(95%<br>CI) | PPV<br>(95%<br>CI) | NPV<br>(95%<br>CI) |
| --- | --- | --- | --- | --- | --- | --- | --- |
| Amylo-Detect | Development<br>(cross-validation) | **0.93**<br>**(0.91;**<br>**0.95)** | **0.93**<br>**(0.92;**<br>**0.93)** | 0.71<br>(0.63;<br>0.79) | **0.93**<br>**(0.92;**<br>**0.93)** | **0.14**<br>**(0.12;**<br>**0.15)** | 1.00<br>(0.99;<br>1.00) |
| Clinical<br>standard | Development<br>(cross-validation) | 0.86<br>(0.84;<br>0.88) | 0.88<br>(0.87;<br>0.88) | **0.84**<br>**(0.80;**<br>**0.89)** | 0.88<br>(0.87;<br>0.89) | 0.13<br>(0.12;<br>0.14) | 1.00<br>(0.99;<br>1.00) |
| Amylo-Detect | Independent<br>internal validation | **0.91**<br>**(0.88;**<br>**0.94)** | **0.93**<br>**(0.92;**<br>**0.94)** | 0.64<br>(0.55;<br>0.74) | **0.94**<br>**(0.93;**<br>**0.95)** | **0.32**<br>**(0.28;**<br>**0.37)** | 0.98<br>(0.98;<br>0.99) |
| Clinical<br>standard | Independent<br>internal validation | 0.84<br>(0.80;<br>0.87) | 0.79<br>(0.78;<br>0.81) | **0.88**<br>**(0.81;**<br>**0.94)** | 0.79<br>(0.77;<br>0.81) | 0.15<br>(0.14;<br>0.17) | **0.99**<br>**(0.99;**<br>**1.00)** |
| Amylo-Detect | External validation | **0.91**<br>**(0.89;**<br>**0.93)** | **0.87**<br>**(0.85;**<br>**0.89)** | 0.73<br>(0.66;<br>0.80) | **0.88**<br>**(0.87;**<br>**0.90)** | **0.39**<br>**(0.35;**<br>**0.44)** | 0.97<br>(0.96;<br>0.98) |
| Clinical<br>standard | External validation | 0.85<br>(0.83;<br>0.87) | 0.75<br>(0.73;<br>0.77) | **0.97**<br>**(0.94;**<br>**0.99)** | 0.73<br>(0.70;<br>0.75) | 0.27<br>(0.26;<br>0.29) | **1.00**<br>**(0.99;**<br>**1.00)** |

AUC, area under the receiver operating characteristic curve; SNS, sensitivity; SPC, specificity; PPV, positive predictive value; NPV, negative predictive value; Bold values indicate higher performance for each comparison.

### Comparison with existing risk score

The *Amylo-Detect* model demonstrated superior performance in the identification of at-risk patients for CA compared to the previously published Mayo ATTR-CM score in the all-comer cohort for all diagnostic metrics, including AUC (all *p*<0.01). In the high pre-test probability cohort with a CMR-ECV >40%, the Mayo ATTR-CM score demonstrated substantially improved performance compared to the all-comer cohort. Nevertheless, the *Amylo-Detect* model continued to outperform the Mayo ATTR-CM score (**Supplemental Table S4**).

### Comparison with current clinical standard

The current clinical standard for the referral of patients for scintigraphy due to the suspicion of CA yielded an AUC of 0.86 (95% CI 0.84; 0.88), 0.84 (95% CI 0.80; 0.87), and 0.85 (95% CI 0.83; 0.87) for cross-validation, internal validation and external validation cohorts respectively. *Amylo-Detect* outperformed the clinical standard in all but 1 or 2 metrics, depending on the cohort (**Table 1**). 10/38 (26%) of the patients missed in clinical routine were identified by *Amylo-Detect*. Incidental detection of high-grade tracer uptake was significantly associated with worse outcome, highlighting the clinical importance of such findings (**Supplemental Figure S5**).

### Subgroup and robustness analyses

Subgroup analyses were performed for the comprehensively annotated Vienna cohorts across prespecified subgroups and demonstrated robust performance (**Figure 2**). This included subgroups where underlying or concomitant CA may be masked by conditions commonly associated with left ventricular thickening. We further performed subgroup analyses for patients with/without carpal tunnel syndrome and available echocardiography data which may sometimes be available after an initial suspicion was raised clinically. In both subgroups, *Amylo-Detect* demonstrated robust performance (AUC 0.88-0.96).

**Figure 2.**
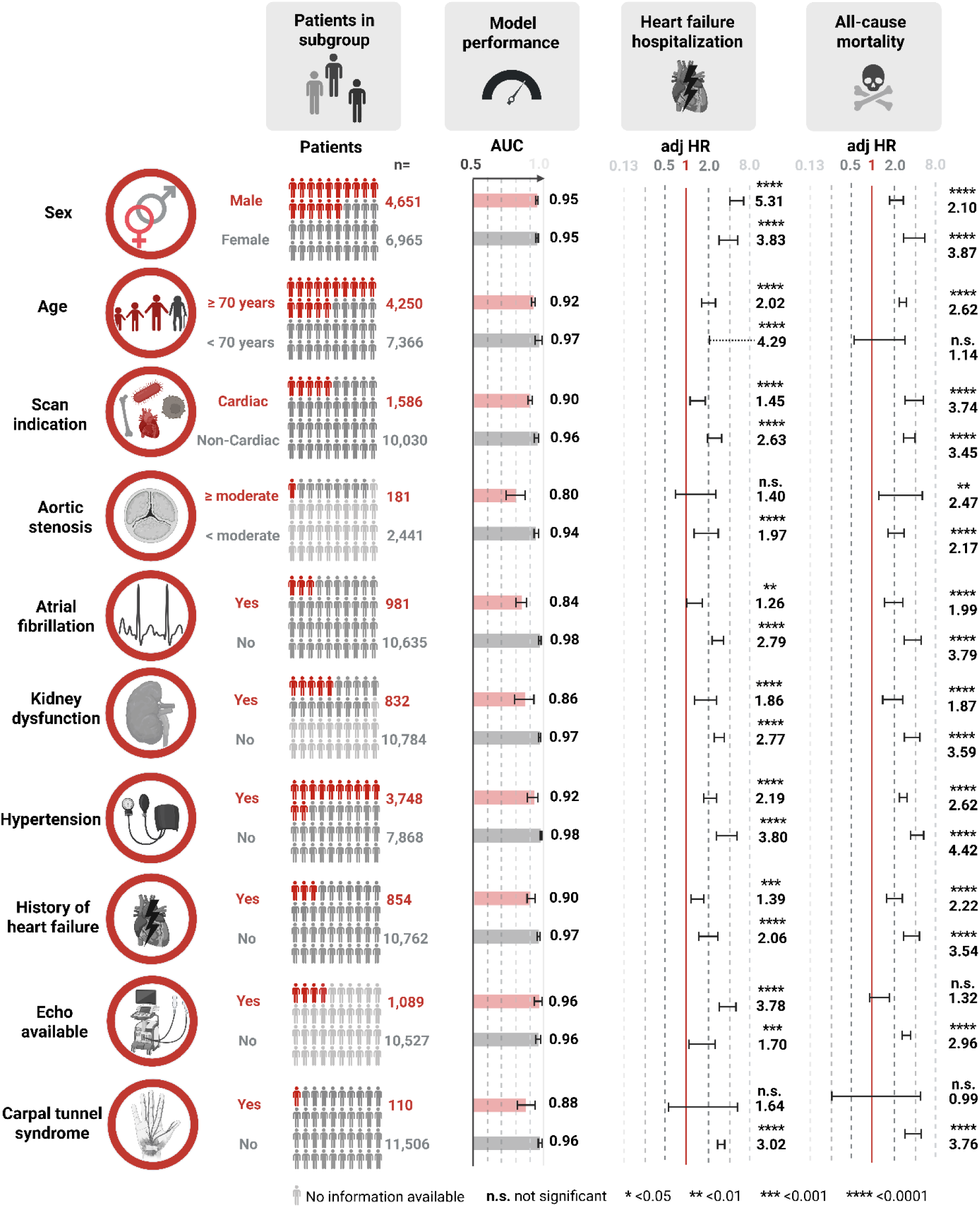
Subgroup analysis results. Cox proportional hazard models for heart failure-associated hospitalization and all-cause mortality were performed based on the prediction by *Amylo-Detect* within the respective subgroups.

On average, patients had 38 missing parameters (IQR 33-46), 6 (IQR 5-7) of which were among the 13 selected parameters in the model. The model demonstrated robustness for missing values and yielded even slightly higher performance for patients with more than 6 missing values, among the selected parameters, compared to patients with fewer missing values (AUC 0.94 [95% CI 0.92; 0.96] vs. AUC 0.91 [95% CI 0.89; 0.93], p<0.05).

We further performed an analysis of patients with high-grade cardac uptake and confirmed CA subtype - either based on biopsy with mass spectrometry or based on single-photon emission computed tomography/computed tomography (SPECT/CT) and light chain assessment. Among the 62 patients included, there were 18 patients with biopsy-confirmed ATTR-CA, 17 (94%) of which were identified as at-risk by *Amylo-Detect*. Of the 3 patients with biopsy-confirmed AL-CA, 2 (67%) were predicted correctly. There were 2 patients with combined ATTR-CA and AL-CA, all (100%) of whom were correctly predicted by *Amylo-Detect*. Among the remaining 39 SPECT-confirmed patients, 31 (79%) were correctly identified.

### Prognostic value

To assess the prognostic value of the model’s predictions, their association with clinical outcomes was evaluated in both the cross-validation and internal validation cohorts (**Figure 3**). After a median follow-up of 66 months (IQR 35-103) after scintigraphy, 590/11,342 (5.2%) patients experienced HFH and 2,877/11,616 (24.8%) patients died. In the cross-validation cohort, predictions were strongly associated with all-cause mortality (adjHR 3.49 [95% CI 3.08; 3.95], p<0.0001) and HFH (adjHR 5.73 [95% CI 4.67; 7.04], p<0.0001). These associations remained significant in the internal validation cohort, both for all-cause mortality (adjHR 2.00 [95% CI 1.42; 2.81], p=0.0007) and HFH (adjHR 4.98 [95% CI 3.18; 7.79], p<0.0001). Among patients not referred for scintigraphy due to CA, incidental high-grade cardiac uptake was significantly associated with all-cause mortality (adjHR 3.47 [95% CI 1.52; 7.92], p<0.0001) and HFH (adjHR 1.70 [95% CI 1.09; 2.66], p=0.02) (**Supplemental Figure S5)**.

**Figure 3.**
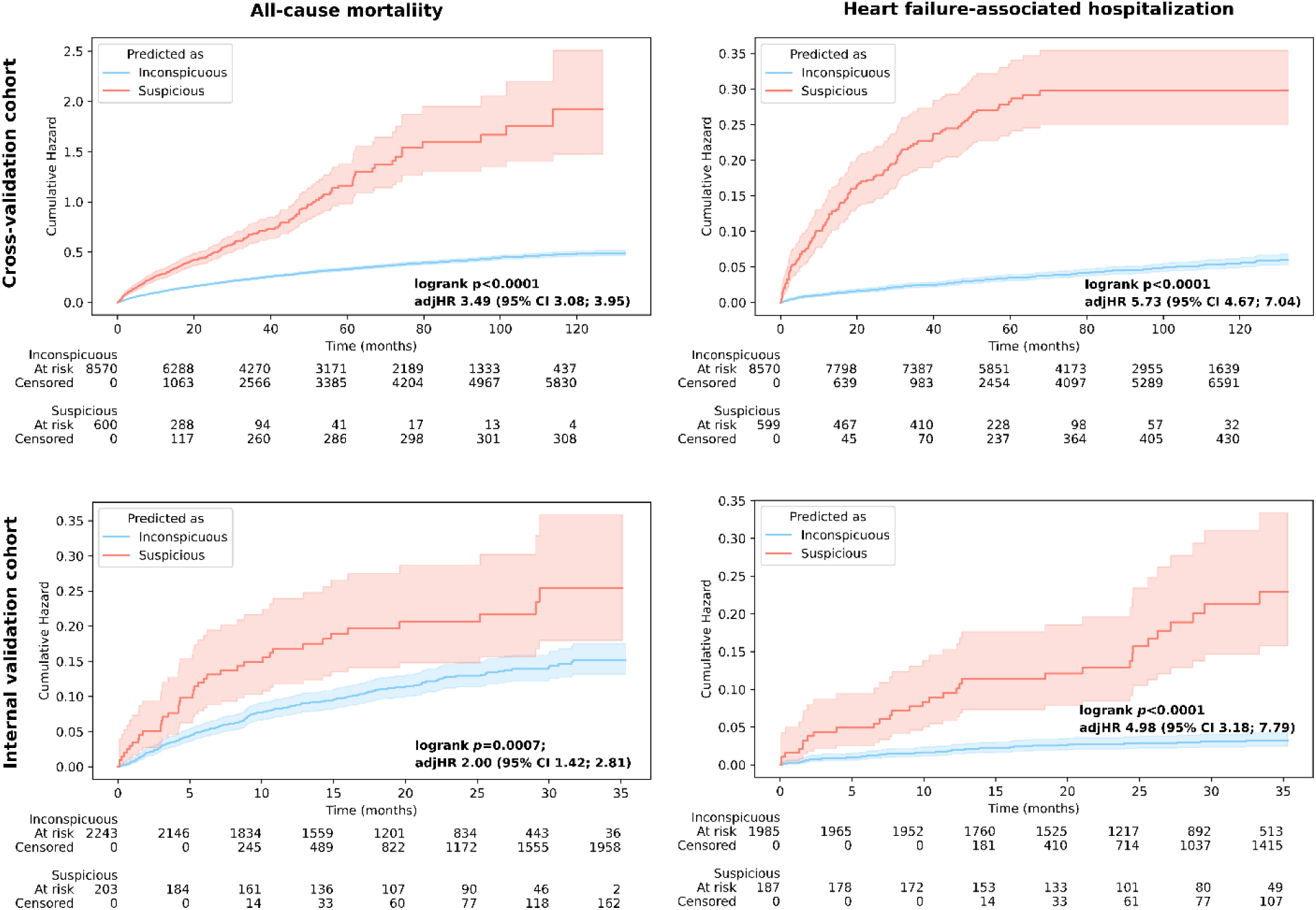
Prognostic value of model predictions. The left column shows Kaplan Meier estimators for all-cause mortality, the right column shows Kaplan Meier estimators for heart failure-associated hospitalization. The upper row shows results for the development (cross-validation) cohort, the lower row shows the results for the independent internal validation.

## Discussion

In this study, we developed and validated *Amylo-Detect*, a prediction tool for identifying patients at risk for CA based on routinely acquired data from EHRs. *Amylo-Detect* demonstrated excellent performance in predicting high-grade cardiac uptake on subsequent bone scans. Among patients not suspected of having CA at the time of scintigraphy, the model correctly predicted CA-suggestive uptake in 26%, highlighting a potential improvement when used in addition to the current clinical referral pathways. The model also conveyed significant prognostic value for HFH and death. We conclude that *Amylo-Detect* may enable fully automated and reliable identification of at-risk patients for CA, prompting confirmatory testing to enable earlier diagnosis and treatment initiation.

CA represents a systemic disease frequently associated with heterogeneous clinical presentation and multiorgan dysfunction^17^. Extracardiac manifestations, such as carpal tunnel syndrome, may precede CA diagnosis by 5-15 years, thereby creating a window of opportunity for early identification while patients are still free from cardiac symptoms^18^. While the presence of extracardiac indicators individually is non-specific, their cumulative presence may raise suspicion for CA. We demonstrate that *Amylo-Detect* can scan EHR and reliably predict the presence of CA-suggestive uptake on subsequent scintigraphy.

Timely treatment initiation is fundamental as novel disease-modifying drugs are available, which yield significant prognostic benefits for affected patients^6–8^. However, respective drugs are less effective at more advanced disease stages^6,8,19^. We hypothesize that machine learning tools, such as *Amylo-Detect*, may shorten the time to diagnosis and identify CA when modern disease-modifying agents exhibit the highest benefit.

Real-world clinical data, particularly from EHR, inherently contains missing values. Accurate integration of this property is crucial to develop reliable and clinically applicable models. Many published models fail to meet this requirement, necessitating additional examination and therefore reducing acceptance and applicability in clinical routine^9,10,20^. In this study, we handle this issue by applying a machine learning-based imputation approach. We show that this approach yields exceptional results over three large-scale cohorts and various clinically relevant subgroups.

A few studies have demonstrated the potential of machine learning to enhance the detection of CA using merely unimodal approaches in late diagnostic settings^21–23^. Three studies have suggested the possibility of integrating structured imaging and non-imaging data for CA identification and referral^9,10,24^. In related studies, Arana-Achaga et al. focused on the detection among ∼1,100 patients with suspected CA while Shiri et al. focused on the detection among ∼260 patients with aortic stenosis and Davies et al. predicted presence of CA in a PYP scan referral cohort, a cohort of HfpEF patients, and an additional external PYP scan referral cohort, including a total of ∼1000 patients. Our study had a 13- to 50-times larger cohort size, included unbiased all-comer referrals for bone scintigraphy, included an external validation on a large-scale cohort, and incorporated prognostic assessment. Unlike the aforementioned studies, this approach enables detection of patients at risk for CA in a real-world setting who would otherwise be missed by mere clinical judgement. *Amylo-Detect* has further demonstrated superior performance with AUCs of 0.94, 0.91, and 0.91 in the development, internal, and external validation cohorts, respectively, compared to AUCs of 0.92 and 0.84 in the study by Arana-Achaga et al.^10^, 0.84 in the study by Shiri et al.^24^, and AUCs of 0.89, 0.84, and 0.84 for the three cohorts employed by Davies et al.^9^. Additionally, in a head-to-head comparison to the Mayo ATTR-CM score^9^ the *Amylo-Detect* model demonstrated substantially improved performance. We further made the developed detection model publicly available via a web-based tool, the *Amylo-Detect* App, to ensure transparency and allow researchers to further validate its use (https://amylo-detect.streamlit.app/).

Users can enter the presence or absence of risk factors and are subsequently provided with a predicted probability of CA-suggestive tracer uptake on bone scintigraphy. The web app allows for tabular input of multiple patients, enabling the usage of *Amylo-Detect* for large-scale studies without the necessity of time-consuming single-patient input.

Several strengths and limitations merit comment. As a systemic disease, many factors may contribute to and facilitate detection in CA. Despite our efforts to include a comprehensive set of parameters, some baseline parameters, such as lumbar spinal stenosis, biceps tendon rupture and electrocardiography data^25–27^ have not been included due to the reliance on structured data from EHR. While our results suggest that *Amylo-Detect* can facilitate earlier diagnosis by detecting at-risk patients before clinical suspicion is raised, it does not necessarily detect early-stage disease. Notably, while *Amylo-Detect* outperformed existing scores as well as the current clinical standard in most scenarios and metrics, it showed lower sensitivity compared to the clinical standard. Nevertheless, *Amylo-Detect* was able to predict a substantial number of cases not identified in clinical routine. Further, the model provides the possibility to adapt the probability threshold to achieve higher sensitivity at the cost of a reduced positive predictive value. In such scenarios, positive predictions by *Amylo-Detect* can be further enhanced by using criteria based on echocardiography and CMR to increase the positive predictive value before patients are sent for final confirmation using scintigraphy and monoclonal protein testing. Development and validation of our model using a broad patient population (all-comer referrals for scintigraphy) ensure real-life applicability. Previous models in the field have been developed using study populations with much higher-than-normal disease prevalence (≥20% vs. 3% in the present cohort), which may yield deceptively good model performance but largely limits their implementation in real-life settings^26,28^. Despite the broad patient population included, it is not feasible to include individuals who did not undergo scintigraphy because of the retrospective study design, as no diagnostic labels would be available for model training. Therefore, our work is subject to a referral bias for bone scintigraphy. Nevertheless, the population spanned a wide range of clinical indications beyond suspected CA and a wide range of ages and comorbidities. This diversity supports the potential generalizability of the model to real-world EHR populations. We further performed rigorous external testing and subgroup analysis, yielding high model performance, even for patients with non-cardiac referral indications who would otherwise have been missed by clinical judgement. This analysis indicates that our results apply to the proposed target population (all patients visiting a hospital). The publication of the *Amylo-Detect* as web app forms the basis for the prospective evaluation of this concept at multiple independent centers.

This study presents a novel machine learning-based approach, *Amylo-Detect*, for the accurate identification of patients at risk for CA, integrating a broad spectrum of available EHR data. *Amylo-Detect* exhibited robust diagnostic performance across multiple cohorts and subgroups, achieving high accuracy for the prediction of high-grade cardiac uptake suggestive for CA, and demonstrated significant prognostic value. *Amylo-Detect* identified many patients with “incidental” CA, not suspected by clinical judgement. This highlights its potential to enhance detection and prompt treatment initiation, crucial for improving clinical outcomes in patients with this progressive and fatal disease.

## Ethics

This study received approval from the Medical University of Vienna’s institutional review board (1557/2020) and was carried out in compliance with the principles of the 1964 Declaration of Helsinki. Owing to the retrospective design of the study, the requirement for informed consent was waived.

## Supporting information

TRUE-AIM Checklist

## Acknowledgments

We want to express our gratitude to the Research Documentation & Analysis (RDA) team of the Medical University of Vienna for their support. We acknowledge BioRender, which was used for the creation of Figure 1 and 2. We thank Yago Lutz for the support in data collection.

## Disclosures

CN reports speaker/consulting honoraria from Pfizer, Bayer, Prothena, and Böhringer Ingelheim and research contracts with Pfizer, AstraZeneca, the Austrian Society of Cardiology, the European Association of Cardiovascular Imaging, and the Austrian Science Fund. CPS received consulting and speaker fees from Pfizer. MH has received lecture fees from Siemens Healthineers and GE Healthcare. MH has received consulting fees from Evomics. TR has received honoraria, lecture fees, and grant support from Edwards Lifesciences, AstraZeneca, Bayer, Novartis, Berlin Chemie, Daiicho-Sankyo, Boehringer Ingelheim, Novo Nordisk, Cardiac Dimensions, and Pfizer. TR is co-founder of mycor GmbH, a company focusing on the development of ECG-algorithms for the identification of amyloidosis. The remaining authors have nothing to disclose.

ChatGPT and Grammarly were used to refine the phrasing of the manuscript; all scientific content and conclusions are the authors’ own.

## Software availability

The developed model is available online via the *Amylo-Detect* App: https://amylo-detect.streamlit.app

## Data availability

The data supporting this study cannot be shared due to restrictions imposed by the ethical approvals. Researchers interested in accessing the data may contact the corresponding author to discuss potential options.

## Author contributors

CPS conceived the study. CPS and CN designed the study. CPS, JY, JH, CN, KK, FH, JN, RC, KM, JM, AK, JM, TTW, CH, and MH collected and annotated the development and internal validation data. DK, KM, SS, JK, TR, and KH collected the external validation data. CN, MH, and CH provided clinical expert opinions and feedback during the study. CPS and DH developed the code. CPS performed the machine learning and statistical analysis. CPS and CN wrote the manuscript and created the figures. All authors critically reviewed the manuscript and provided feedback.

## References

1. Garcia-Pavia, P. et al. Diagnosis and treatment of cardiac amyloidosis. A position statement of the European Society of Cardiology Working Group on Myocardial and Pericardial Diseases. Eur. J. Heart Fail. 23, 512–526 (2021).

2. González-López, E. et al. Wild-type transthyretin amyloidosis as a cause of heart failure with preserved ejection fraction. Eur. Heart J. 36, 2585–2594 (2015).

3. AbouEzzeddine, O. F. et al. Prevalence of Transthyretin Amyloid Cardiomyopathy in Heart Failure With Preserved Ejection Fraction. JAMA Cardiol 6, 1267–1274 (2021).

4. Nitsche, C. et al. Prevalence and Outcomes of Concomitant Aortic Stenosis and Cardiac Amyloidosis. J. Am. Coll. Cardiol. 77, 128–139 (2021).

5. Nitsche, C. et al. Prevalence and outcomes of cardiac amyloidosis in all-comer referrals for bone scintigraphy. J. Nucl. Med. (2022) doi:10.2967/jnumed.122.264041.

6. Maurer, M. S. et al. Tafamidis Treatment for Patients with Transthyretin Amyloid Cardiomyopathy. N. Engl. J. Med. 379, 1007–1016 (2018).

7. Gillmore, J. D. et al. Efficacy and Safety of Acoramidis in Transthyretin Amyloid Cardiomyopathy. N. Engl. J. Med. 390, 132–142 (2024).

8. Fontana, M. et al. Vutrisiran in patients with transthyretin amyloidosis with cardiomyopathy. N. Engl. J. Med. (2024) doi:10.1056/NEJMoa2409134.

9. Davies, D. R. et al. A Simple Score to Identify Increased Risk of Transthyretin Amyloid Cardiomyopathy in Heart Failure With Preserved Ejection Fraction. JAMA Cardiol 7, 1036–1044 (2022).

10. Arana-Achaga, X. et al. Development and validation of a prediction model and score for transthyretin cardiac amyloidosis diagnosis: T-Amylo. JACC Cardiovasc. Imaging 16, 1567–1580 (2023).

11. Peng, H., Long, F. & Ding, C. Feature selection based on mutual information: criteria of max-dependency, max-relevance, and min-redundancy. IEEE Trans. Pattern Anal. Mach. Intell. 27, 1226–1238 (2005).

12. Chawla, N. V., Bowyer, K. W., Hall, L. O. & Kegelmeyer, W. P. SMOTE: Synthetic Minority Over-sampling Technique. jair 16, 321–357 (2002).

13. Garcia-Pavia, P. et al. Diagnosis and treatment of cardiac amyloidosis: a position statement of the ESC Working Group on Myocardial and Pericardial Diseases. Eur. Heart J. 42, 1554–1568 (2021).

14. Kidoh, M. et al. Cardiac MRI-derived extracellular volume fraction versus myocardium-to-lumen R1 ratio at postcontrast T1 mapping for detecting cardiac amyloidosis. Radiol. Cardiothorac. Imaging 5, e220327 (2023).

15. Kitaoka, H. et al. JCS 2020 guideline on diagnosis and treatment of cardiac amyloidosis. Circ. J. 84, 1610–1671 (2020).

16. Kolossváry, M. et al. The transparent reporting and evaluation of artificial intelligence models (TRUE-AIM) report card: A quality assurance initiative of the American heart association journals to promote high-quality, reproducible artificial intelligence research. Circulation 152, e311–e322 (2025).

17. Ioannou, A. et al. Multiorgan dysfunction and associated prognosis in transthyretin cardiac amyloidosis. J. Am. Heart Assoc. 13, e033094 (2024).

18. Westin, O. et al. Screening for cardiac amyloidosis 5 to 15 years after surgery for bilateral carpal tunnel syndrome. J. Am. Coll. Cardiol. 80, 967–977 (2022).

19. Elliott, P., Gundapaneni, B., Sultan, M. B., Ines, M. & Garcia-Pavia, P. Improved long-term survival with tafamidis treatment in patients with transthyretin amyloid cardiomyopathy and severe heart failure symptoms. Eur. J. Heart Fail. 25, 2060–2064 (2023).

20. Hong, Z. et al. Enhanced diagnostic and prognostic assessment of cardiac amyloidosis using combined 11C-PiB PET/CT and 99mTc-DPD scintigraphy. Eur. J. Nucl. Med. Mol. Imaging 1–12 (2025) doi:10.1007/s00259-025-07157-7.

21. Spielvogel, C. P. et al. Diagnosis and prognosis of abnormal cardiac scintigraphy uptake suggestive of cardiac amyloidosis using artificial intelligence: a retrospective, international, multicentre, cross-tracer development and validation study. Lancet Digit Health 6, e251–e260 (2024).

22. Huda, A. et al. A machine learning model for identifying patients at risk for wild-type transthyretin amyloid cardiomyopathy. Nat. Commun. 12, 2725 (2021).

23. Halme, H.-L. et al. Convolutional neural networks for detection of transthyretin amyloidosis in 2D scintigraphy images. EJNMMI Res. 12, 27 (2022).

24. Shiri, I. et al. Multi-modality artificial intelligence-based transthyretin amyloid cardiomyopathy detection in patients with severe aortic stenosis. Eur. J. Nucl. Med. Mol. Imaging 1–16 (2024) doi:10.1007/s00259-024-06922-4.

25. Geller, H. I., Singh, A., Alexander, K. M., Mirto, T. M. & Falk, R. H. Association between ruptured distal biceps tendon and wild-type transthyretin cardiac amyloidosis. JAMA 318, 962–963 (2017).

26. Oikonomou, E. K. et al. Artificial intelligence-enabled electrocardiography and echocardiography to track preclinical progression of transthyretin amyloid cardiomyopathy. Eur. Heart J. (2025) doi:10.1093/eurheartj/ehaf450.

27. Youmans, Q. R., Shah, S. J. & Khan, S. S. Hereditary transthyretin amyloid cardiomyopathy. JAMA Cardiol. 7, 236 (2022).

28. Slivnick, J. A. et al. Cardiac amyloidosis detection from a single echocardiographic video clip: a novel artificial intelligence-based screening tool. Eur. Heart J. 46, 4090–4101 (2025).

