## Supplemental Material for "Screening for patients at risk for cardiac amyloidosis via electronic health records: A multicenter machine learning development and validation study"

**Table S1.** Clinical characteristics of the development cohort.

| Parameter | Parameter value | *p* value | Entire data | No high-grade uptake | High-grade uptake |
| --- | --- | --- | --- | --- | --- |
| Number of patients, n (%) |  |  | 9,170 (100.00%) | 9,028 (98.45%) | 142 (1.55%) |
| High-grade cardiac uptake (Perugini grade 2 or 3) |  | < 0.0001 | 142 (1.55%) | 0 (0%) | 142 (100.0%) |
| Age (years) |  | < 0.0001 | 61.77 (15.73) | 61.5 (15.67) | 78.68 (8.41) |
| BMI (kg/m^2^) |  | 0.2606 | 26.38 (5.32) | 26.39 (5.33) | 25.71 (4.27) |
|  | NA |  | 277 (3.02%) | 274 (3.04%) | 3 (2.11%) |
| Male sex |  | < 0.0001 | 3,326 (36.27%) | 3,218 (35.64%) | 108 (76.05%) |
| **Comorbidities** | | | | | |
| Atrial fibrillation |  | < 0.0001 | 857 (9.35%) | 786 (8.71%) | 71 (50.0%) |
| History of any bleeding event |  | 0.8483 | 481 (5.25%) | 473 (5.24%) | 8 (5.63%) |
| History of cerebral bleeding |  | 0.3858 | 86 (0.94%) | 84 (0.93%) | 2 (1.41%) |
| Carpal tunnel syndrome |  | < 0.0001 | 62 (0.68%) | 34 (0.38%) | 28 (19.72%) |
| COPD |  | 0.1363 | 657 (7.16%) | 642 (7.11%) | 15 (10.56%) |
| Coronary artery disease |  | < 0.0001 | 1,201 (13.1%) | 1,162 (12.87%) | 39 (27.46%) |
| History of myocardial infarction |  | 0.1973 | 174 (1.9%) | 169 (1.87%) | 5 (3.52%) |
| Diabetes |  | 0.7973 | 1,131 (12.33%) | 1,115 (12.35%) | 16 (11.27%) |
| History of heart failure |  | < 0.0001 | 828 (9.03%) | 767 (8.5%) | 61 (42.96%) |
| Hyperlipidemia |  | 0.5967 | 250 (2.73%) | 248 (2.75%) | 2 (1.41%) |
| Hypertension |  | 0.1209 | 3,723 (40.6%) | 3,656 (40.5%) | 67 (47.18%) |
| Liver disease |  | 0.0227 | 418 (4.56%) | 417 (4.62%) | 1 (0.7%) |
| Myeloma |  | 1.0000 | 93 (1.01%) | 92 (1.02%) | 1 (0.7%) |
| Renal insufficiency (ICD-10) |  | < 0.0001 | 784 (8.55%) | 755 (8.36%) | 29 (20.42%) |
| History of stroke |  | 0.0382 | 507 (5.53%) | 493 (5.46%) | 14 (9.86%) |
| **Blood parameters** | | | | | |
| NT-proBNP (pg/mL) |  | < 0.0001 | 2,848 (5,292) | 2,700 (5,220) | 5,227 (6,123) |
|  | NA |  | 7,488 (81.66%) | 7,450 (82.52%) | 38 (26.76%) |
| Creatinine (µmol/L) |  | < 0.0001 | 1.05 (0.8) | 1.04 (0.8) | 1.34 (0.77) |
|  | NA |  | 4,034 (43.99%) | 4,012 (44.44%) | 22 (15.49%) |
| eGFR (mL/min/1.73 m²) |  | < 0.0001 | 80.14 (31.12) | 80.57 (31.17) | 62.13 (22.55) |
|  | NA |  | 4,034 (43.99%) | 4,012 (44.44%) | 22 (15.49%) |
| BUN (mg/dL) |  | < 0.0001 | 17.84 (10.39) | 17.6 (10.09) | 27.67 (16.09) |
|  | NA |  | 4,090 (44.60%) | 4,068 (45.06%) | 22 (15.49%) |
| Triglycerides (mg/dL) |  | < 0.0001 | 131.15 (74.61) | 132.21 (75.01) | 90.05 (38.8) |
|  | NA |  | 5,376 (58.63%) | 5,330 (59.04%) | 46 (32.39%) |
| Cholesterol (mg/dL) |  | < 0.0001 | 183.12 (52.01) | 183.81 (52.01) | 157.01 (44.74) |
|  | NA |  | 5,422 (59.13%) | 5,376 (59.55%) | 46 (32.39%) |
| LDL (mg/dL) |  | < 0.0001 | 105.08 (43.35) | 105.86 (43.36) | 82.27 (36.21) |
|  | NA |  | 6,995 (76.28%) | 6,925 (76.71%) | 70 (49.30%) |
| Bilirubin (mg/dL) |  | < 0.0001 | 0.68 (1.13) | 0.68 (1.14) | 0.85 (0.58) |
|  | NA |  | 4,488 (48.94%) | 4,461 (49.41%) | 27 (19.01%) |
| Hemoglobin (g/dL) |  | 0.0348 | 12.61 (1.93) | 12.6 (1.94) | 12.98 (1.72) |
|  | NA |  | 4,209 (45.90%) | 4,190 (46.41%) | 19 (13.38%) |
| Hematocrit (%) |  | 0.0199 | 37.46 (5.41) | 37.43 (5.42) | 38.61 (4.85) |
|  | NA |  | 4,209 (45.90%) | 4,190 (46.41%) | 19 (13.38%) |
| HbA1c (%) |  | 0.0583 | 5.92 (1.11) | 5.92 (1.13) | 5.95 (0.73) |
|  | NA |  | 7,529 (82.10%) | 7,450 (82.52%) | 79 (55.63%) |
| TSH (mIU/L) |  | 0.8108 | 2.16 (3.02) | 2.14 (2.9) | 2.64 (5.74) |
|  | NA |  | 6,319 (68.91%) | 6,257 (69.31%) | 62 (43.66%) |
| LDH (U/L) |  | < 0.0001 | 230.02 (178.64) | 229.64 (179.89) | 246.61 (111.07) |
|  | NA |  | 4,607 (50.24%) | 4,569 (50.61%) | 38 (26.76%) |
| Troponin (ng/L) |  | < 0.0001 | 101.82 (507.68) | 101.01 (519.86) | 116.17 (192.15) |
|  | NA |  | 8,164 (89.03%) | 8,076 (89.46%) | 88 (61.97%) |
| Creatine kinase (U/L) |  | 0.0502 | 178.83 (436.13) | 179.41 (440.94) | 158.65 (203.31) |
|  | NA |  | 5,806 (63.32%) | 5,757 (63.77%) | 49 (34.51%) |
| Creatine kinase MB (U/L) |  | 0.7851 | 69.31 (164.79) | 70.37 (167.22) | 41.16 (69.1) |
|  | NA |  | 8,645 (94.27%) | 8,522 (94.40%) | 123 (86.62%) |
| CRP (mg/L) |  | 0.0179 | 3.45 (6.27) | 3.47 (6.28) | 2.44 (5.76) |
|  | NA |  | 4,389 (47.86%) | 4,360 (48.29%) | 29 (20.42%) |
| **Echocardiography** | | | | | |
| Left ventricular end-diastolic diameter (mm) |  | 0.0024 | 44.81 (16.37) | 44.94 (16.61) | 41.33 (6.12) |
|  | NA |  | 7,879 (85.92%) | 7,782 (86.20%) | 97 (68.31%) |
| Left atrial diameter (mm) |  | < 0.0001 | 54.13 (9.18) | 53.82 (9.07) | 62.39 (8.27) |
|  | NA |  | 7,939 (86.58%) | 7,841 (86.85%) | 98 (69.01%) |
| Right ventricular end-diastolic diameter (mm) |  | 0.0817 | 31.9 (8.2) | 31.84 (8.26) | 33.57 (6.24) |
|  | NA |  | 7,888 (86.02%) | 7,792 (86.31%) | 96 (67.61%) |
| Right atrial diameter (mm) |  | < 0.0001 | 52.61 (8.64) | 52.31 (8.5) | 60.47 (8.46) |
|  | NA |  | 7,894 (86.09%) | 7,799 (86.39%) | 95 (66.90%) |
| IVS thickness (mm) |  | < 0.0001 | 14.14 (5.19) | 13.9 (5.08) | 20.07 (4.29) |
|  | NA |  | 8,012 (87.37%) | 7,914 (87.66%) | 98 (69.01%) |
| Tricuspid regurgitation | < moderate | < 0.0001 | 1,749 (19.07%) | 1,704 (18.88%) | 45 (31.03%) |
|  | ≥ moderate |  | 215 (2.34%) | 189 (2.09%) | 26 (17.93%) |
|  | NA |  | 7,206 (78.58%) | 7,132 (79.02%) | 74 (51.03%) |
| Pulmonary regurgitation | < moderate | 1.0000 | 1,957 (21.34%) | 1,886 (20.9%) | 71 (48.97%) |
|  | ≥ moderate |  | 7 (0.08%) | 7 (0.08%) | 0 (0%) |
|  | NA |  | 7,206 (78.58%) | 7,132 (79.02%) | 74 (51.03%) |
| Mitral regurgitation | < moderate | < 0.0001 | 1,782 (19.43%) | 1,731 (19.18%) | 51 (35.17%) |
|  | ≥ moderate |  | 182 (1.98%) | 162 (1.8%) | 20 (13.79%) |
|  | NA |  | 7,206 (78.58%) | 7,132 (79.02%) | 74 (51.03%) |
| Mitral stenosis | < moderate | 1.0000 | 1,962 (21.4%) | 1,891 (20.95%) | 71 (48.97%) |
|  | ≥ moderate |  | 2 (0.02%) | 2 (0.02%) | 0 (0%) |
|  | NA |  | 7,206 (78.58%) | 7,132 (79.02%) | 74 (51.03%) |
| Aortic regurgitation | < moderate | 0.5600 | 1,927 (21.01%) | 1,857 (20.58%) | 70 (48.28%) |
|  | ≥ moderate |  | 22 (0.24%) | 21 (0.23%) | 1 (0.69%) |
|  | NA |  | 7,221 (78.75%) | 7,147 (79.19%) | 74 (51.03%) |
| Aortic stenosis | < moderate | 0.6395 | 1,822 (19.87%) | 1,757 (19.47%) | 65 (44.83%) |
|  | ≥ moderate |  | 142 (1.55%) | 136 (1.51%) | 6 (4.14%) |
|  | NA |  | 7,206 (78.58%) | 7,132 (79.02%) | 74 (51.03%) |
| Left ventricular function (diastolic) | Normal | < 0.0001 | 131 (1.43%) | 131 (1.45%) | 0 (0%) |
|  | Grade 1 dysfunction |  | 584 (6.37%) | 578 (6.4%) | 6 (4.23%) |
|  | Grade 2 dysfunction |  | 115 (1.25%) | 112 (1.24%) | 3 (2.11%) |
|  | Grade 3 dysfunction |  | 13 (0.14%) | 8 (0.09%) | 5 (3.52%) |
|  | Grade 4 dysfunction |  | 48 (0.52%) | 39 (0.43%) | 9 (6.34%) |
|  | NA |  | 8,279 (90.28%) | 8,160 (90.39%) | 119 (83.8%) |
| Left ventricular function (systolic) | Preserved | < 0.0001 | 1,072 (11.69%) | 1,051 (11.64%) | 21 (14.79%) |
|  | Mildly reduced |  | 110 (1.52%) | 128 (1.42%) | 12 (8.45%) |
|  | Moderately reduced |  | 59 (0.65%) | 54 (0.6%) | 5 (3.52%) |
|  | Severely reduced |  | 68 (0.74%) | 51 (0.68%) | 7 (4.93%) |
|  | NA |  | 7,831 (85.40%) | 7,734 (85.67%) | 97 (68.31%) |
| **Cardiac MRI** | | | | | |
| Left ventricular ejection fraction (%) |  | 0.0765 | 56.57 (15.66) | 57.1 (15.65) | 51.51 (14.83) |
|  | NA |  | 8,948 (97.58%) | 8,827 (97.77%) | 121 (85.21%) |
| Right ventricular ejection fraction (%) |  | 0.0739 | 51.76 (12.32) | 52.21 (12.18) | 47.5 (12.8) |
|  | NA |  | 8,949 (97.59%) | 8,828 (97.78%) | 121 (85.21%) |
| Right ventricular end-diastolic diameter (mm) (mm) |  | 0.8670 | 39.61 (7.27) | 39.54 (6.99) | 40.24 (9.4) |
|  | NA |  | 8,954 (97.64%) | 8,833 (97.84%) | 121 (85.21%) |
| Left ventricular end-diastolic volume (mL) |  | 0.1473 | 155.76 (53.62) | 154.04 (52.96) | 172.1 (57.01) |
|  | NA |  | 8,950 (97.60%) | 8,829 (97.80%) | 121 (85.21%) |
| Right ventricular end-diastolic volume (mL) |  | 0.0401 | 152.19 (51.08) | 149.75 (50.01) | 175.24 (55.23) |
|  | NA |  | 8,950 (97.60%) | 8,829 (97.80%) | 121 (85.21%) |
| Left ventricular end-systolic volume (mL) |  | 0.0435 | 71.65 (45.55) | 69.74 (44.59) | 89.81 (50.25) |
|  | NA |  | 8,950 (97.60%) | 8,829 (97.80%) | 121 (85.21%) |
| Left ventricular stroke volume (mL) |  | 0.9684 | 84.18 (26.56) | 84.37 (27.07) | 82.38 (21.1) |
|  | NA |  | 8,950 (97.60%) | 8,829 (97.80%) | 121 (85.21%) |
| Right ventricular stroke volume (mL) |  | 0.5604 | 76.88 (25.59) | 76.68 (26.08) | 78.81 (20.3) |
|  | NA |  | 8,950 (97.60%) | 8,829 (97.80%) | 121 (85.21%) |
| Left ventricular cardiac output (L/min) |  | 0.6797 | 5.52 (1.66) | 5.53 (1.71) | 5.51 (1.03) |
|  | NA |  | 8,950 (97.60%) | 8,829 (97.80%) | 121 (85.21%) |
| Right ventricular cardiac output (L/min) |  | 0.1508 | 5.05 (1.65) | 5.02 (1.7) | 5.3 (1.14) |
|  | NA |  | 8,950 (97.60%) | 8,829 (97.80%) | 121 (85.21%) |
| Left ventricular cardiac index (L/min/m²) |  | 0.4452 | 2.95 (0.93) | 2.94 (0.96) | 3.02 (0.52) |
|  | NA |  | 9,008 (98.23%) | 8,880 (98.36%) | 128 (90.14%) |
| Right ventricular cardiac index (L/min/m²) |  | 0.1373 | 2.67 (0.87) | 2.65 (0.89) | 2.9 (0.59) |
|  | NA |  | 9,004 (98.19%) | 8,876 (98.32%) | 128 (90.14%) |
| Left ventricular end-systolic diameter (mm) |  | 0.5331 | 47.35 (7.29) | 47.45 (7.11) | 46.45 (8.68) |
|  | NA |  | 8,951 (97.61%) | 8,831 (97.82%) | 120 (84.51%) |
| Left ventricular end-diastolic diameter (mm) |  | 0.5331 | 47.35 (7.29) | 47.45 (7.11) | 46.45 (8.68) |
|  | NA |  | 8,951 (97.61%) | 8,831 (97.82%) | 120 (84.51%) |
| Extracellular volume (%) |  | < 0.0001 | 30.05 (8.61) | 28.83 (6.99) | 43.78 (12.33) |
|  | NA |  | 8,987 (98.00%) | 8,860 (98.14%) | 127 (89.44%) |

Continuous parameters are given as mean and standard deviation. Binary and ordinal parameters are given as absolute numbers and prevalence. BMI, body mass index; COPD, chronic obstructive pulmonary disease; NT-proBNP, N-terminal pro B-type natriuretic peptide; eGFR, estimated glomerular filtration rate; BUN, blood urea nitrogen; LDL, low-density lipoprotein; HbA1c, Hemoglobin A1c; TSH, thyroid stimulating hormone; LDH, lactate dehydrogenase; CRP, C-reactive protein.

**Table S2.** Clinical characteristics of the independent internal validation cohort.

| Parameter | Parameter value | *p* value | Entire data | No high-grade uptake | High-grade uptake |
| --- | --- | --- | --- | --- | --- |
| Number of patients, n (%) |  |  | 2,446 (100.00%) | 2,345 (95.87%) | 101 (4.13%) |
| High-grade cardiac uptake (Perugini grade 2 or 3) |  | < 0.0001 | 101 (4.13%) | 0 (0%) | 101 (100.0%) |
| Age (years) |  | < 0.0001 | 64.08 (15.91) | 63.43 (15.84) | 79.2 (8.13) |
| BMI (kg/m^2^) |  | 0.4362 | 27.12 (5.19) | 27.07 (5.17) | 28.96 (5.36) |
|  | NA |  | 2,201 (89.98%) | 2,106 (89.81%) | 95 (94.06%) |
| Male sex |  | < 0.0001 | 1,325 (54.17%) | 1,242 (52.96%) | 83 (82.18%) |
| **Comorbidities** | | | | | |
| Atrial fibrillation |  | 0.0004 | 124 (5.07%) | 110 (4.69%) | 14 (13.86%) |
| History of any bleeding event |  | 0.7209 | 49 (2.0%) | 48 (2.05%) | 1 (0.99%) |
| History of cerebral bleeding |  | < 0.0001 | 35 (1.43%) | 10 (0.43%) | 25 (24.75%) |
| Carpal tunnel syndrome |  | 0.4476 | 48 (1.96%) | 45 (1.92%) | 3 (2.97%) |
| COPD |  | 1 | 34 (1.39%) | 33 (1.41%) | 1 (0.99%) |
| Coronary artery disease |  | 0.0277 | 118 (4.82%) | 108 (4.61%) | 10 (9.9%) |
| History of myocardial infarction |  | 1 | 39 (1.59%) | 38 (1.62%) | 1 (0.99%) |
| Diabetes |  | < 0.0001 | 145 (5.93%) | 126 (5.37%) | 19 (18.81%) |
| History of heart failure |  | 0.2919 | 26 (1.06%) | 24 (1.02%) | 2 (1.98%) |
| Hyperlipidemia |  | 0.4633 | 112 (4.58%) | 106 (4.52%) | 6 (5.94%) |
| Hypertension |  | 0.6229 | 25 (1.02%) | 25 (1.07%) | 0 (0%) |
| Liver disease | NA |  | 2,446 (100.0%) | 2,345 (100.0%) | 101 (100.0%) |
| Myeloma |  | 0.6640 | 37 (1.51%) | 35 (1.49%) | 2 (1.98%) |
| Renal insufficiency (ICD-10) |  | 0.0456 | 48 (1.96%) | 43 (1.83%) | 5 (4.95%) |
| History of stroke |  | 0.0366 | 31 (1.27%) | 27 (1.15%) | 4 (3.96%) |
| **Blood parameters** | | | | | |
| NT-proBNP (pg/mL) |  | < 0.0001 | 2,066 (5,322) | 1,873 (5,217) | 4,481 (5,991) |
|  | NA |  | 1,378 (56.34%) | 1,356 (57.83%) | 22 (21.78%) |
| Creatinine (µmol/L) |  | < 0.0001 | 1.05 (0.82) | 1.04 (0.82) | 1.36 (0.65) |
|  | NA |  | 580 (23.71%) | 569 (24.26%) | 11 (10.89%) |
| eGFR (mL/min/1.73 m²) | NA |  | 2,446 (100.00%) | 2,345 (100.00%) | 101 (100.00%) |
|  |  | < 0.0001 | 18.43 (10.8) | 17.9 (10.05) | 28.86 (17.55) |
| BUN (mg/dL) | NA |  | 595 (24.33%) | 584 (24.90%) | 11 (10.89%) |
|  |  | < 0.0001 | 133.37 (82.91) | 135.22 (84.11) | 97.25 (39.9) |
| Triglycerides (mg/dL) | NA |  | 891 (36.43%) | 866 (36.93%) | 25 (24.75%) |
|  |  | < 0.0001 | 175.82 (53.37) | 177.28 (53.3) | 147.45 (46.35) |
| Cholesterol (mg/dL) | NA |  | 929 (37.98%) | 902 (38.46%) | 27 (26.73%) |
| LDL (mg/dL) | NA |  | 2,446 (100.00%) | 2,345 (100.00%) | 101 (100.00%) |
|  |  | < 0.0001 | 0.56 (0.52) | 0.54 (0.51) | 0.85 (0.62) |
| Bilirubin (mg/dL) | NA |  | 642 (26.25%) | 631 (26.91%) | 11 (10.89%) |
|  |  | 0.2326 | 12.92 (1.92) | 12.91 (1.92) | 13.19 (2.05) |
| Hemoglobin (g/dL) | NA |  | 604 (24.69%) | 593 (25.29%) | 11 (10.89%) |
|  |  | 0.1003 | 38.67 (5.35) | 38.62 (5.34) | 39.61 (5.63) |
| Hematocrit (%) | NA |  | 604 (24.69%) | 593 (25.29%) | 11 (10.89%) |
|  |  | 0.0176 | 5.95 (1.11) | 5.95 (1.14) | 5.95 (0.72) |
| HbA1c (%) | NA |  | 1,496 (61.16%) | 1,459 (62.22%) | 37 (36.63%) |
|  |  | 0.4724 | 2.08 (2.46) | 2.09 (2.51) | 1.84 (1.27) |
| TSH (mIU/L) | NA |  | 1,191 (48.69%) | 1,156 (49.30%) | 35 (34.65%) |
|  |  | < 0.0001 | 223.21 (121.62) | 222.52 (123.97) | 235.9 (63.28) |
| LDH (U/L) | NA |  | 1,034 (42.27%) | 1,006 (42.90%) | 28 (27.72%) |
|  | NA |  | 2,446 (100.00%) | 2,345 (100.00%) | 101 (100.00%) |
| Troponin (ng/L) |  | 0.0438 | 109.5 (119.79) | 108.81 (120.98) | 120.57 (98.21) |
|  | NA |  | 1,260 (51.51%) | 1,229 (52.41%) | 31 (30.69%) |
| Creatine kinase (U/L) |  | 0.9721 | 28.77 (67.39) | 29.16 (69.3) | 22.31 (12.58) |
|  | NA |  | 1,935 (79.11%) | 1,863 (79.45%) | 72 (71.29%) |
| Creatine kinase MB (U/L) |  | 0.6977 | 1.47 (3.37) | 1.49 (3.39) | 1.2 (3.01) |
|  | NA |  | 993 (40.60%) | 972 (41.45%) | 21 (20.79%) |
| **Echocardiography** | | | | | |
| Left ventricular end-diastolic diameter (mm) |  | < 0.0001 | 44.21 (6.4) | 44.59 (6.31) | 39.62 (5.64) |
|  | NA |  | 1,958 (80.05%) | 1,894 (80.77%) | 64 (63.37%) |
| Left atrial diameter (mm) |  | 0.0345 | 53.18 (9.42) | 52.87 (9.43) | 58.08 (7.7) |
|  | NA |  | 2,242 (91.66%) | 2,153 (91.81%) | 89 (88.12%) |
| Right ventricular end-diastolic diameter (mm) |  | 0.8965 | 32.41 (5.21) | 32.4 (5.23) | 32.46 (4.92) |
|  | NA |  | 1,956 (79.97%) | 1,892 (80.68%) | 64 (63.37%) |
| Right atrial diameter (mm) |  | < 0.0001 | 53.94 (9.45) | 53.53 (9.45) | 58.92 (7.87) |
|  | NA |  | 1,953 (79.84%) | 1,889 (80.55%) | 64 (63.37%) |
| IVS thickness (mm) |  | < 0.0001 | 15.05 (3.99) | 14.61 (3.68) | 20.46 (3.71) |
|  | NA |  | 1,982 (81.03%) | 1,916 (81.71%) | 66 (65.35%) |
| Tricuspid regurgitation | < moderate | 0.0008 | 572 (23.39%) | 537 (22.87%) | 35 (35.71%) |
|  | ≥ moderate |  | 86 (3.52%) | 71 (3.02%) | 15 (15.31%) |
|  | NA |  | 1,788 (73.1%) | 1,740 (74.11%) | 48 (48.98%) |
| Pulmonary regurgitation | < moderate | 1.0000 | 655 (26.78%) | 605 (25.77%) | 50 (51.02%) |
|  | ≥ moderate |  | 3 (0.12%) | 3 (0.13%) | 0 (0%) |
|  | NA |  | 2,446 (100.00%) | 2,345 (100.00%) | 101 (100.00%) |
| Mitral regurgitation | < moderate | 0.0036 | 571 (23.34%) | 535 (22.79%) | 36 (36.73%) |
|  | ≥ moderate |  | 87 (3.56%) | 73 (3.11%) | 14 (14.29%) |
|  | NA |  | 1,788 (73.1%) | 1,740 (74.11%) | 48 (48.98%) |
| Mitral stenosis | < moderate | 1.0000 | 658 (26.9%) | 608 (25.89%) | 50 (51.02%) |
|  | ≥ moderate |  | 0 (0.00%) | 0 (0.00%) | 0 (0.00%) |
|  | NA |  | 1,788 (73.1%) | 1,740 (74.11%) | 48 (48.98%) |
| Aortic regurgitation | < moderate | 1.0000 | 635 (25.96%) | 587 (25.0%) | 48 (48.98%) |
|  | ≥ moderate |  | 13 (0.53%) | 12 (0.51%) | 1 (1.02%) |
|  | NA |  | 1,798 (73.51%) | 1,749 (74.49%) | 49 (50.0%) |
| Aortic stenosis | < moderate | 0.0638 | 619 (25.31%) | 569 (24.23%) | 50 (51.02%) |
|  | ≥ moderate |  | 39 (1.59%) | 39 (1.66%) | 0 (0%) |
|  | NA |  | 1,788 (73.1%) | 1,740 (74.11%) | 48 (48.98%) |
| Left ventricular function (diastolic) | Normal | < 0.0001 | 30 (1.23%) | 30 (1.28%) | 0 (0%) |
|  | Grade 1 dysfunction |  | 190 (7.77%) | 185 (7.89%) | 5 (4.95%) |
|  | Grade 2 dysfunction |  | 50 (2.04%) | 46 (1.96%) | 4 (3.96%) |
|  | Grade 3 dysfunction |  | 8 (0.33%) | 5 (0.21%) | 3 (2.97%) |
|  | Grade 4 dysfunction |  | 11 (0.45%) | 10 (0.43%) | 1 (0.99%) |
|  | NA |  | 2,157 (88.18%) | 2,069 (88.23%) | 88 (87.13%) |
| Left ventricular function (systolic) | Preserved | < 0.0001 | 352 (14.40%) | 338 (14.42%) | 14 (13.86%) |
|  | Mildly reduced |  | 75 (3.06%) | 62 (2.64%) | 13 (12.87%) |
|  | Moderately reduced |  | 42 (1.71%) | 34 (1.45%) | 8 (8.92%) |
|  | Severely reduced |  | 26 (1.07%) | 24 (1.02%) | 2 (1.98%) |
|  | NA |  | 1,951 (79.76%) | 1,887 (80.47%) | 64 (63.37%) |
| **Cardiac MRI** | | | | | |
| Left ventricular ejection fraction (%) |  | 0.0607 | 54.15 (14.41) | 55.6 (14.89) | 48.79 (10.91) |
|  | NA |  | 2,380 (97.30%) | 2,293 (97.78%) | 87 (86.14%) |
| Right ventricular ejection fraction (%) |  | 0.3764 | 50.85 (12.0) | 51.72 (11.87) | 47.86 (11.97) |
|  | NA |  | 2,384 (97.47%) | 2,297 (97.95%) | 87 (86.14%) |
| Right ventricular end-diastolic diameter (mm) (mm) |  | 0.2882 | 40.71 (6.44) | 41.0 (6.19) | 39.79 (7.11) |
|  | NA |  | 2,387 (97.59%) | 2,300 (98.08%) | 87 (86.14%) |
| Left ventricular end-diastolic volume (mL) |  | 0.5572 | 1,64.75 (52.54) | 164.39 (56.19) | 166.0 (36.97) |
|  | NA |  | 2,383 (97.42%) | 2,296 (97.91%) | 87 (86.14%) |
| Right ventricular end-diastolic volume (mL) |  | 0.3014 | 168.12 (67.56) | 166.21 (71.72) | 174.79 (49.81) |
|  | NA |  | 2,383 (97.42%) | 2,296 (97.91%) | 87 (86.14%) |
| Left ventricular end-systolic volume (mL) |  | 0.1261 | 78.16 (43.31) | 75.92 (46.72) | 86.0 (26.84) |
|  | NA |  | 2,383 (97.42%) | 2,296 (97.91%) | 87 (86.14%) |
| Left ventricular stroke volume (mL) |  | 0.3407 | 86.97 (26.01) | 89.11 (26.74) | 79.93 (22.03) |
|  | NA |  | 2,386 (97.55%) | 2,299 (98.04%) | 87 (86.14%) |
| Right ventricular stroke volume (mL) |  | 0.8888 | 83.75 (27.76) | 84.35 (28.03) | 81.79 (26.74) |
|  | NA |  | 2,386 (97.55%) | 2,299 (98.04%) | 87 (86.14%) |
| Left ventricular cardiac output (L/min) |  | 0.5758 | 5.57 (1.45) | 5.69 (1.47) | 5.19 (1.31) |
|  | NA |  | 2,386 (97.55%) | 2,299 (98.04%) | 87 (86.14%) |
| Right ventricular cardiac output (L/min) |  | 0.8749 | 5.37 (1.53) | 5.36 (1.35) | 5.41 (1.99) |
|  | NA |  | 2,386 (97.55%) | 2,299 (98.04%) | 87 (86.14%) |
| Left ventricular cardiac index (L/min/m²) |  | 0.3467 | 2.94 (0.74) | 3.02 (0.77) | 2.71 (0.58) |
|  | NA |  | 2,395 (97.91%) | 2,308 (98.42%) | 87 (86.14%) |
| Right ventricular cardiac index (L/min/m²) |  | 0.7465 | 2.83 (0.73) | 2.83 (0.65) | 2.83 (0.92) |
|  | NA |  | 2,393 (97.83%) | 2,306 (98.34%) | 87 (86.14%) |
| Left ventricular end-systolic diameter (mm) |  | 0.1053 | 47.57 (6.7) | 48.17 (6.7) | 45.57 (6.3) |
|  | NA |  | 2,386 (97.55%) | 2,299 (98.04%) | 87 (86.14%) |
| Left ventricular end-diastolic diameter (mm) |  | 0.1053 | 47.57 (6.7) | 48.17 (6.7) | 45.57 (6.3) |
|  | NA |  | 2,386 (97.55%) | 2,299 (98.04%) | 87 (86.14%) |
| Extracellular volume (%) |  | 0.5604 | 76.88 (25.59) | 76.68 (26.08) | 78.81 (20.3) |
|  | NA |  | 8,950 (97.60%) | 8,829 (97.80%) | 121 (85.21%) |
| Left ventricular ejection fraction (%) |  | 0.6797 | 5.52 (1.66) | 5.53 (1.71) | 5.51 (1.03) |
|  | NA |  | 8,950 (97.60%) | 8,829 (97.80%) | 121 (85.21%) |
| Right ventricular ejection fraction (%) |  | 0.1508 | 5.05 (1.65) | 5.02 (1.7) | 5.3 (1.14) |
|  | NA |  | 8,950 (97.60%) | 8,829 (97.80%) | 121 (85.21%) |
| Right ventricular end-diastolic diameter (mm) (mm) |  | 0.4452 | 2.95 (0.93) | 2.94 (0.96) | 3.02 (0.52) |
|  | NA |  | 9,008 (98.23%) | 8,880 (98.36%) | 128 (90.14%) |
| Left ventricular end-diastolic volume (mL) |  | 0.1373 | 2.67 (0.87) | 2.65 (0.89) | 2.9 (0.59) |
|  | NA |  | 9,004 (98.19%) | 8,876 (98.32%) | 128 (90.14%) |
| Right ventricular end-diastolic volume (mL) |  | 0.5331 | 47.35 (7.29) | 47.45 (7.11) | 46.45 (8.68) |
|  | NA |  | 8,951 (97.61%) | 8,831 (97.82%) | 120 (84.51%) |
| Left ventricular end-systolic volume (mL) |  | 0.5331 | 47.35 (7.29) | 47.45 (7.11) | 46.45 (8.68) |
|  | NA |  | 8,951 (97.61%) | 8,831 (97.82%) | 120 (84.51%) |
| Left ventricular stroke volume (mL) |  | < 0.0001 | 30.05 (8.61) | 28.83 (6.99) | 43.78 (12.33) |
|  | NA |  | 8,987 (98.00%) | 8,860 (98.14%) | 127 (89.44%) |

Continuous parameters are given as mean and standard deviation. Binary and ordinal parameters are given as absolute numbers and prevalence. BMI, body mass index; COPD, chronic obstructive pulmonary disease; NT-proBNP, N-terminal pro B-type natriuretic peptide; eGFR, estimated glomerular filtration rate; BUN, blood urea nitrogen; LDL, low-density lipoprotein; HbA1c, Hemoglobin A1c; TSH, thyroid stimulating hormone; LDH, lactate dehydrogenase; CRP, C-reactive protein.

**Table S3.** Clinical characteristics of the external validation cohort.

| Parameter | Parameter value | *p* value | Entire data | No high-grade uptake | High-grade uptake |
| --- | --- | --- | --- | --- | --- |
| Number of patients, n (%) |  |  | 1521 (100.00%) | 1376 (90.47%) | 145 (9.53%) |
| High-grade cardiac uptake (Perugini grade 2 or 3) |  | < 0.0001 | 145 (9.53%) | 0 (0%) | 145 (100.0%) |
| Male sex |  | < 0.0001 | 639 (42.01%) | 517 (37.57%) | 122 (84.14%) |
|  | NA |  | 9 (0.59%) | 7 (0.51%) | 2 (1.38%) |
| Carpal tunnel syndrome |  | < 0.0001 | 110 (7.23%) | 45 (3.27%) | 65 (44.83%) |
| Atrial fibrillation |  | < 0.0001 | 374 (24.59%) | 270 (19.62%) | 104 (71.72%) |
|  | NA |  | 23 (1.51%) | 21 (1.53%) | 2 (1.38%) |
| Interventricular septum thickness (mm) |  | < 0.0001 | 13.52 (4.53) | 12.79 (4.1) | 17.64 (4.62) |
|  | NA |  | 1143 (75.15%) | 1055 (76.67%) | 88 (60.69%) |
| Left ventricular function (systolic) | Preserved | 0.00984 | 190 (12.49%) | 172 (12.5%) | 18 (12.41%) |
|  | Mildly reduced |  | 67 (4.4%) | 54 (3.92%) | 13 (8.97%) |
|  | Moderately reduced |  | 74 (4.87%) | 57 (4.14%) | 17 (11.72%) |
|  | Severely reduced |  | 10 (0.66%) | 10 (0.73%) | 0 (0%) |
|  | NA |  | 1180 (77.58%) | 1083 (78.71%) | 97 (66.9%) |

Continuous parameters are given as mean and standard deviation. Binary and ordinal parameters are given as absolute numbers and prevalence.

**Table S4.** Performance comparison of Amylo-Detect with existing risk score (Mayo ATTR-CM). Performance of the superior model for each metric and analysis is shown in bold.

| Model | Cohort | Subgroup | AUC | SNS | SPC | PPV | NPV |
| --- | --- | --- | --- | --- | --- | --- | --- |
| Amylo-Detect | Cross-validation | All | **0.93**  **(0.91; 0.95)** | **0.71 (0.63; 0.79)** | **0.93 (0.92; 0.93)** | **0.14 (0.12; 0.15)** | **1.00 (0.99; 1.00)** |
| Davies et al. score 2022 | Cross-validation | All | 0.55  (0.51; 0.59) | 0.50  (0.42; 0.58) | 0.60  (0.59; 0.61) | 0.02  (0.02; 0.02) | 0.99  (0.99; 0.99) |
| Amylo-Detect | Independent validation | All | **0.91 (0.88; 0.94)** | 0.64 (0.55; 0.74) | **0.94 (0.93; 0.95)** | **0.32 (0.28; 0.37)** | **0.98 (0.98; 0.99)** |
| Davies et al. score 2022 | Independent validation | All | 0.59  (0.54; 0.64) | 0.64  (0.55; 0.73) | 0.54 (0.52; 0.56) | 0.06 (0.05; 0.07) | 0.97 (0.97; 0.98) |
| Amylo-Detect | Cross-validation | ECV >40% (n=16) | **0.84**  **(0.64; 1.00)** | **1.00**  **(1.00; 1.00)** | 0.43  (0.14; 0.86) | **0.69**  **(0.60; 0.90)** | **1.00**  **(1.00; 1.00)** |
| Davies et al. score 2022 | Cross-validation | ECV >40% (n=16) | 0.72  (0.62; 0.82) | 0.48  (0.29; 0.67) | **0.97**  **(0.95; 0.98)** | 0.40  (0.24; 0.57) | 0.98  (0.97; 0.99) |
| Amylo-Detect | Independent validation | ECV >40% (n=14) | **1.00**  **(1.00; 1.00)** | **1.00**  **(1.00; 1.00)** | NA | **0.79**  **(0.79; 0.79)** | NA |
| Davies et al. score 2022 | Independent validation | ECV >40% (n=14) | 0.83  (0.69; 0.95) | 0.75  (0.50; 1.00) | 0.91  (0.82; 0.98) | 0.69  (0.50; 0.92) | 0.93  (0.87; 1.00) |

Better performance in each comparison is displayed in bold. AUC, area under the receiver operating characteristic curve; SNS, sensitivity; SPC, specificity; PPV, positive predictive value; NPV, negative predictive value; NA, not applicable (all patients predicted to be at-risk).

**Figure S1.** Referral indications for patients in the development (cross-validation) and independent internal validation data from Vienna.

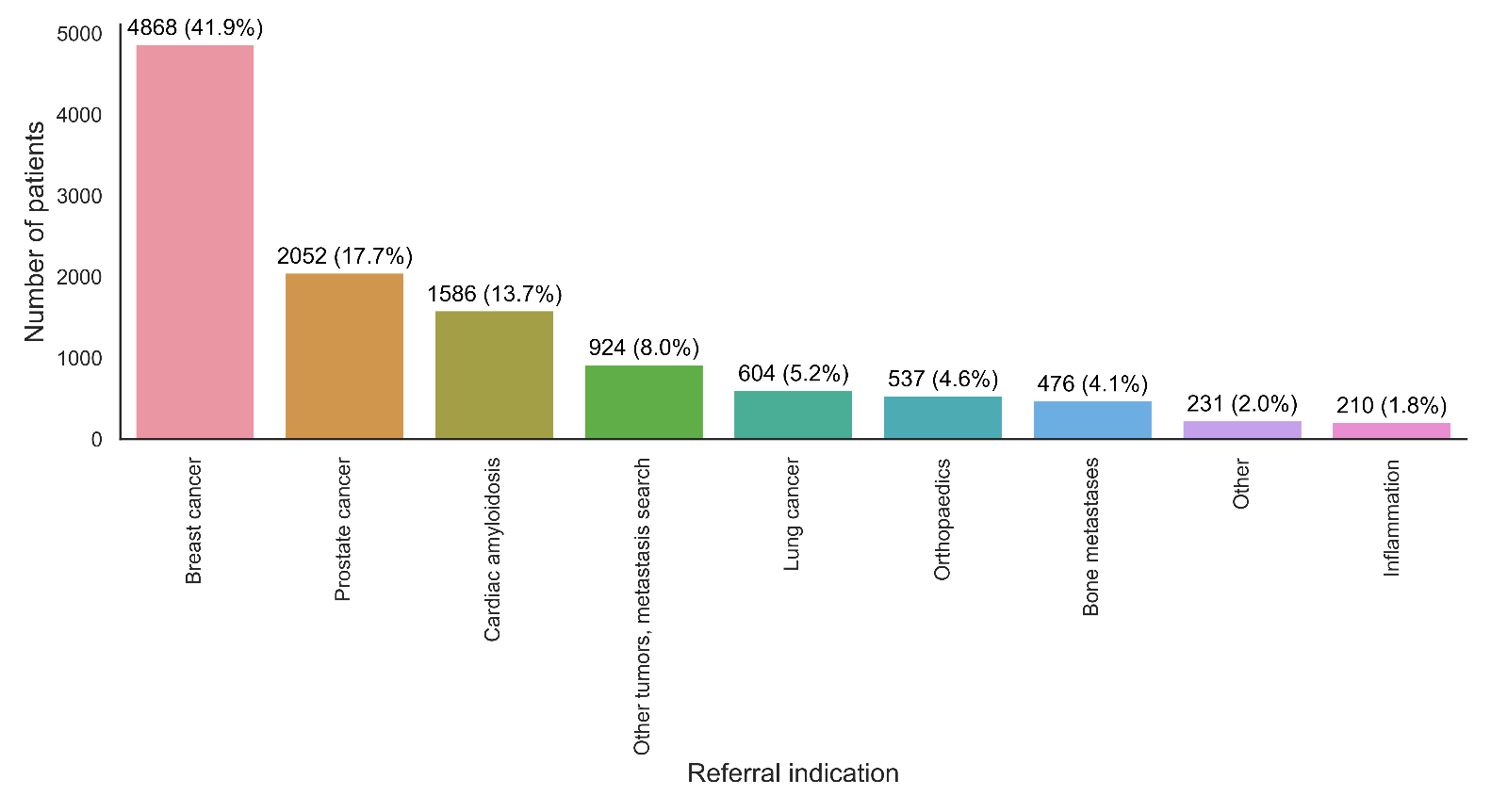

**Figure S2.** Cohort flow diagram.

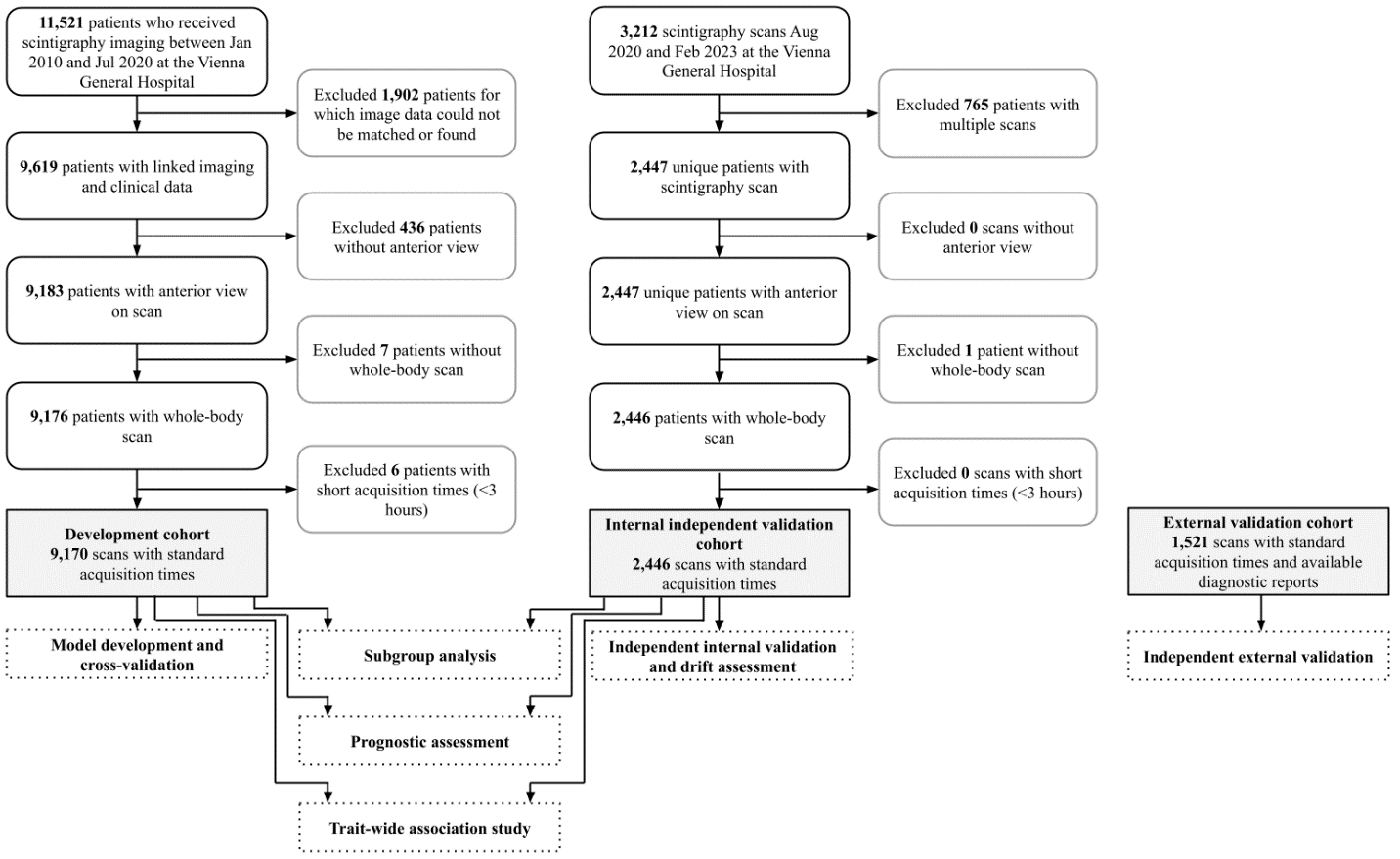

**Figure S3.** Performance for multiple evaluated algorithms during development in the cross-validation cohort.

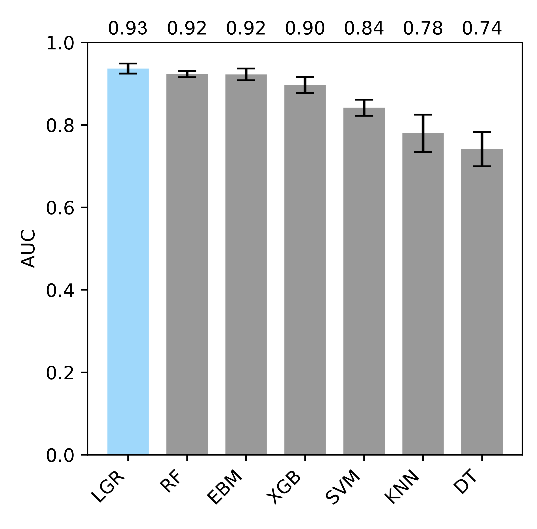

Area under the receiver operating characteristic curve (AUC) for the logistic regression model which was used for subsequent analyses is shown in blue. Values above the bars indicate AUC values for the corresponding algorithm. LGR, Machine learning-enhanced logistic regression; RF, Random forest; EBM, Explainable boosting machine; XGB, Extreme gradient boosting; SVM, Support vector machine; KNN, k-nearest neighbors; DT, Decision tree; AUC, Area under the curve.

**Figure S4.** Risk calibration curves for the high-uptake model before and after calibration using isotonic regression.

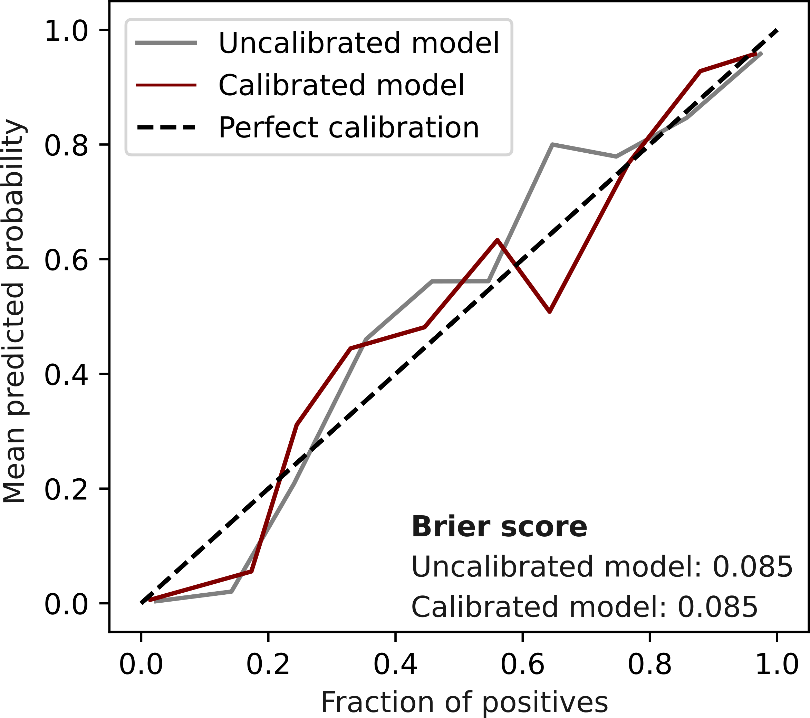

Coefficients of the logistic regression models indicating the relative importance and direction of the association (red indicates positive association) with the prediction target are shown for high cardiac uptake (C). Higher values for ‘Left ventricular function’ indicate worse function.

**Figure S5.** Prognostic value of high-grade cardiac uptake in patients not suspected of cardiac amyloidosis at the time of scintigraphy among patients in the two Vienna cohorts.

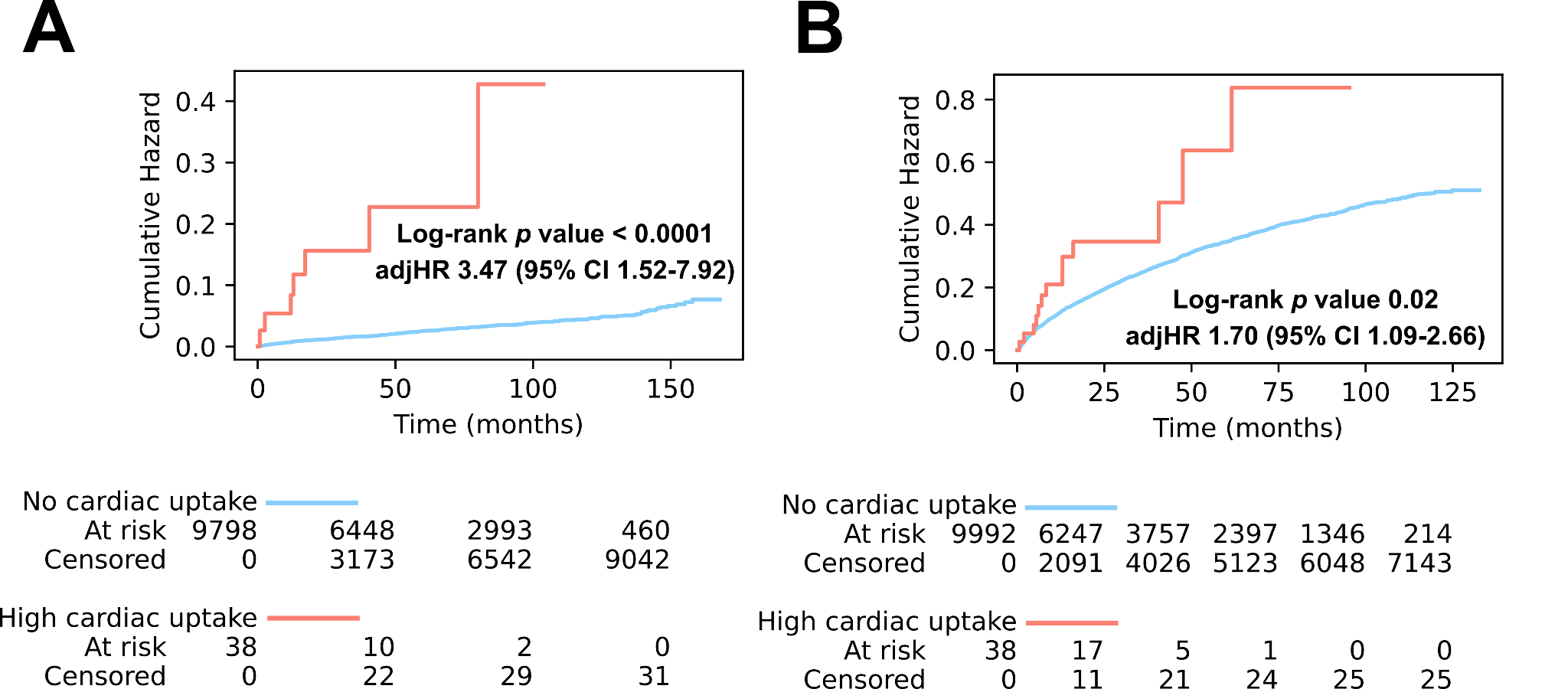

**A)** Heart failure hospitalization. **B)** All-cause mortality. Red Kaplan Meier estimators indicate patients with high-grade cardiac uptake (Perugini grade 2 or 3). Blue Kaplan Meier estimators indicate patients with Perugini grade 0 or 1.
