## Supplementary material for "Screening for patients at risk for cardiac amyloidosis via electronic health records: A multicenter machine learning development and validation study": TRUE-AIM Checklist

**How to complete this report card:**

Response column: Please provide a brief answer to the question. If the cell includes predefined single-choice answers, choose the most appropriate answer and delete the other options.

Comment column: Please provide further explanation or context, if needed

Page No column: Provide the page number where the given claim is discussed in the clean manuscript file

Please complete all required questions (shaded in green) and additional questions as indicated by the answers to the required questions.

**To be completed for all manuscripts that use AI as part of the Methods**

| **Study design** | | | | | |
| --- | --- | --- | --- | --- | --- |
| **ID.** | **Topic** | **Description** | **Response** | **Comment** | **Page no.** |
| 1 | Primary objective | What is the primary objective of the current paper? | This study aimed to develop and validate a machine learning model, Amylo-Detect, using structured multimodal electronic health record (EHR) data to guide referrals for confirmatory scintigraphy and monoclonal protein testing. |  | 3 |
| 2 | Primary outcome | What is the primary outcome variable of the current project? | Diagnostic performance in the detection of high grade cardiac uptake suggestive for cardiac amyloidosis |  | 3 |
| 3 | Software | What software and corresponding packages (with version numbers) used for given parts of the analysis? | All developments and analyses were performed using Python 3.9.5. |  | 8 |
| 4 | Random seed | Was the random seed number set to a specific value prior to all analyses? | Yes | Prior to all analyses, random seeds were set to the default where available, otherwise to zero. | 8 |

**To be completed for all manuscripts that use AI as part of the Methods**

**Data**

| **ID.** | **Topic** | **Description** | **Response** | **Comment** | **Page no.** |
| --- | --- | --- | --- | --- | --- |
| **Training set** | | | | | |
| 101 | Training set acquisition | How was data acquired for the training set? | Retrospectively (all-comers for bone scintigraphy due to any indication) |  | 5 |
| 102 | Training set sites | How many different sites contributed to the training set? | One |  |  |
| 103 | Training set characteristics | What was the sample size, proportion of race/sex/ethnicity groups and the distribution of outcomes in the training data? | All subgroups and corresponding proportions are shown in Supplemental Table 1. Race and ethnicity are not collected at the participating centers and therefore not reported. |  | Suppl 2-5 |
| 104 | Training set overlap | Are multiple instances of a sample present in the training data? | No |  | 8 |
| 105 | Training set availability | Are all deidentified training data (tabulated, images, etc.) acquired for study purposes of the training set needed to replicate the results available? | No | For replication, we provide a simple-to-use web app | 7 |
| **Validation Set** (Only answer 201 to 207 if the answer to question 200 is *Yes* or *Partially)* | | | | | |
| 200 | Validation set | Was a validation set (separate to the test set) used during the development? | No |  |  |
| 201 | Validation set acquisition | How was data acquired for the validation set? | - |  |  |
| 202 | Validation set sites | How many different sites contributed to the validation set? | - |  |  |
| 203 | Validation set characteristics | What was the sample size, proportion of race/sex/ethnicity groups and the distribution of outcomes in the validation data? | - |  |  |
| 204 | Validation set overlap | Are multiple instances of a sample present in the validation data? | - |  |  |
| 205 | Validation set availability | Are all deidentified validation data (tabulated, images, etc.) acquired for study purposes of the validation set needed to replicate the results available? | - |  |  |
| 206 | Validation and training set overlap | Were there instances of the same sample in both in the training and validation set? | - |  |  |
| 207 | Validation and training set separation | How were the training and validation set separated from each other? | - |  |  |
| **Test Set** (Only answer 301 to 308 if the answer to question 300 is *Yes* or *Partially)* | | | | | |
| 300 | Test set | Does the analysis have a separate test set? | Yes |  | 5 |
| 301 | Test set acquisition | How was data acquired for the test set? | Retrospectively (all-comers for bone scintigraphy) |  | 5 |
| 302 | Test set sites | How many different sites contributed to the test set? | Two |  | 5 |
| 303 | Test set characteristics | What was the sample size, proportion of race/sex/ethnicity groups and the distribution of outcomes in the test data? | All subgroups and corresponding proportions are shown in Supplemental Tables 2 and 3. Race and ethnicity are not collected at the participating centers and therefore not reported. |  | Suppl 6-10 |
| 304 | Test set overlap | Are multiple instances of a sample present in the test data? | No |  | 8 |
| 305 | Test set availability | Are all deidentified test data (tabulated, images, etc.) acquired for study purposes of the test set needed to replicate the results available? | May be made available upon request if a dedicated ethical approval is in place, allowing sharing of the data | Data supporting this study cannot be shared due to restrictions imposed by the ethical approvals. Researchers interested in accessing the data may contact the corresponding author to discuss potential options. | 18 |
| 306 | Test and training set overlap | Were there instances of the same sample in both in the training and test set? | No |  | 6-7 |
| 307 | Test and training set separation | How were the training and test set separated from each other? | Cross-validation for development cohort, independent split for internal independent validation (= test set), independent split for external independent validation (= external test set) |  | 6 |
| 308 | Test set type | Are the results from the test set considered internal or external evaluation? | Both | We performed three separate evaluations of the model, including an internal and an external evaluation | 6 |
| **Data characteristics** | | | | | |
| 400 | Missing data | What is the type of missingness in the training, validation and test data? | Missing at random |  | 6 |
| 401 | Strategy for missing data | How was missingness handled in the analysis? | Imputation | Imputation was exclusively fitted on the training data | 6 |
| 402 | Class imbalance | Was the data imbalanced requiring resampling or other measures? | Yes |  | 6 |
| 403 | Strategy for class imbalance | How was imbalance handled? | Synthetic minority oversampling technique (SMOTE), | Fitted solely on the training data | 6 |

**To be completed if applicable to the manuscript**

| **ID.** | **Topic** | **Description** | **Response** | **Comment** | **Page no.** |
| --- | --- | --- | --- | --- | --- |
| **A) Data generation and augmentation** (Only answer A1 to A4 if the answer to question A is *Yes using AI*, *Yes without AI*, or *Partially*) | | | | | |
| A | Data generation and augmentation | Were new instances created, or data augmented? | No |  |  |
| A1 | Generated data availability | Are all newly created instances by the generative model available? | - |  |  |
| A2 | Bias mitigation | Were measures taken to determine whether the generated dataset was representative and free from bias? | - |  |  |
| A3 | Train-time augmentation | Was data augmentation done during any part of model development? | - |  |  |
| A4 | Code availability | Is all code used for data generation and/or augmentation purposes available? | - |  |  |
| **B) Preprocessing** (Only answer B1 to B9 if the answer to question B is *Yes using AI*, *Yes without AI*, or *Partially*) | | | | | |
| B | Preprocessing | Were preprocessing steps done prior to analysis? | Yes |  | 6 |
| B1 | Feature scaling | What type of feature scaling was applied prior to analysis? | Standardization | Solely fitted on the training data | 6 |
| B2 | Image preprocessing | What preprocessing steps were done prior to image analysis? | Standardization, imputation, selection of features, addressing imbalances with SMOTE, automated hyperparameter tuning | All solely fitted on the training data | 6 |
| B3 | Collinearity assessment | Were actions taken to address multicollinearity of input features? | No |  |  |
| B4 | Feature selection | Were features filtered prior to evaluation? | Supervised | Using mRMR, solely fitted on the training data | 6 |
| B5 | Dimensionality reduction | Were dimensionality reduction techniques used to reduce the number of features? | No |  |  |
| B6 | Hyperparameters | How were the hyperparameters of the preprocessing pipeline optimized? | Minimum redundancy maximum relevance (mRMR) | Solely fitted on the training data | 6 |
| B7 | Reproducibility | Was reproducibility of the preprocessing pipeline evaluated? | Yes | All preprocessing is fully reproducible using the publicly available web app | 7 |
| B8 | Preprocessing dataset | Were instances used to evaluate model performance part of the dataset used to develop the preprocessing pipeline? | No |  | 6 |
| B9 | Code availability | Is all code used for preprocessing purposes available? | Publicly available | The web app employed processing and inference are available via the web app | 7 |
| **C) Feature generation** (Only answer C1 to C6 if the answer to question C is *Yes using AI*, *Yes without AI*, or *Partially*) | | | | | |
| C | Feature generation | Were new features created? | No |  |  |
| C1 | Methodology | What method was used for generating new features? | - |  |  |
| C2 | Optimization | On which data was the feature generation conducted? | - |  |  |
| C3 | Hyperparameters | How were the hyperparameters of the feature generation models optimized? | - |  |  |
| C4 | Reproducibility | Was reproducibility of new generated features evaluated? | - |  |  |
| C5 | Feature generation dataset overlap | Were instances used to evaluate model performance part of the dataset used for feature development? | - |  |  |
| C6 | Code availability | Is all code used for feature generation purposes available? | - |  |  |
| **D) Model development** (Only answer D1 to D11 if the answer to question D is *Yes using AI*, *Yes without AI*, or *Partially*) | | | | | |
| D | Model development | Were statistical models created? | Yes | Diagnostic models using AI and Prognostic models without AI | 8 |
| D1 | Model initiation | Was a pretrained model used to initiate values of the model? | No |  |  |
| D2 | Methodology | What method was used for model building? | Supervised |  | 6-7 |
| D3 | Optimization | On which data were the model parameters (including cut-off values) optimized? | Training data |  | 6-7 |
| D4 | Hyperparameters | How were the hyperparameters of the models optimized? | Grid search algorithm |  | 6 |
| D5 | Test-time augmentation and ensemble techniques | Were methods used to further improve the prediction performance? | Aforementioned preprocessing methods (solely fitted on training data) |  | 6-7 |
| D6 | Overfitting | How was potential overfitting addressed? | Aforementioned preprocessing methods (solely fitted on training data), evaluation on independent internal and external test cohorts |  | 5 |
| D7 | Bias mitigation | How were potential model biases handled? | Comprehensive subgroup analysis for evaluation of biases |  | 11 |
| D8 | Calibration | Were models calibrated? | No | Calibration was evaluated but did not show an improvement over non-calibrated models in terms of Brier score | Suppl 15 |
| D9 | Model building dataset | Were instances used to evaluate model performance part of the dataset used to develop the models? | No |  | 6 |
| D10 | Code availability | Is all code used for model building purposes available? | Publicly available | With this publication, we supply a publicly available web app containing all code for inference and reproduction | 7 |
| D11 | Model availability | Is the fully trained model available? | Yes, publicly available |  | 7 |
| **E) Model evaluation** (Only answer E1 to E8 if the answer to question E is *Yes using AI*, *Yes without AI*, or *Partially*) | | | | | |
| E | Model evaluation | Were model performances evaluated and compared? | Yes |  | 6 |
| E1 | Evaluation | On what data were the models evaluated? | Models were evaluated on three different cohorts: 1) Via cross-validation on the development cohort 2) Independent cohort (from same center as development cohort but a different time range) 3) from an independent external center |  | 6 |
| E2 | Performance metrics | What performance metrics were used to evaluate model performance? | Comprehensive diagnostic metrics, prognostic metrics for Cox models | Diagnostic: AUC, accuracy, sensitivity, specificity, positive predictive value, negative predictive value  Prognostic: adjusted hazard ratios | 12, 23 |
| E3 | Model comparisons | What statistical tests were used to compare models? | DeLong |  |  |
| E4 | Performance uncertainty | Was the uncertainty of model performance evaluated? | Yes | 95% confidence intervals are provided for all scenarios and metrics | throughout |
| E5 | Sub-group analysis | Were model biases evaluated (performance reported in different age and sex race / ethnicity groups)? | Yes |  | 11 |
| E6 | Superiority of generated features | Were models including newly generated features compared to more simple models including clinical and/or quantitative features readily available? | Yes | We compared to an existing score and the clinical standard | 10 |
| E7 | Superiority of machine learning models | Was the performance of machine learning models compared to more simple statistical models (e.g. regression)? | Yes | Existing score | 10 |
| E8 | Code availability | Is all code used for model evaluation purposes available? | Publicly available | Via web app | 7 |
| **F) Model interpretation** (Only answer F1 to F4 if the answer to question F is *Yes using AI*, *Yes without AI*, or *Partially*) | | | | | |
| F | Model interpretation | Were models used to interpret results? | No |  |  |
| F1 | Methodology | What methods were used to help interpretation of model reasoning? | - |  |  |
| F2 | Hyperparameters | How were the hyperparameters of the models optimized? | - |  |  |
| F3 | Multiple methods | Were multiple methods used to interpret the results of the models? | - |  |  |
| F4 | Code availability | Is all code used for model interpretation purposes available? | - |  |  |

Posted 10/8/2025
